# Non-invasive technology to assess hydration status in advanced cancer to explore relationships between fluid status and symptoms: an observational study using bioelectrical impedance analysis

**DOI:** 10.1101/2023.08.25.23294216

**Authors:** Amara Callistus Nwosu, Sarah Stanley, Catriona R Mayland, Stephen Mason, Alexandra McDougall, John E Ellershaw

## Abstract

**Background:** Oral fluid intake decreases in people with advanced cancer, especially when they approach the dying phase of their illness. There is inadequate evidence to support hydration assessment and decision making in the dying phase of illness. Bioelectrical impedance analysis (BIA) and vector analysis (BIVA) are validated methods of hydration assessment, with research demonstrating that hydration status is associated with specific symptoms, and survival in advanced cancer. However, further research is needed to better understand the relationships between hydration status and clinical outcomes in advanced cancer, particularly at the end-of-life.

**Aim:** To evaluate hydration status and its associations with clinical outcomes in advanced cancer patients, and those in the last week of life.

**Materials and methods:** An observational study of people with advanced cancer in three centres. Advance consent methodology was used to conduct hydration assessments in the dying. Total body water was estimated using the BIA Impedance index (Height – H (m)^2^/Resistance – R (Ohms)). We used backward regression to identify factors (signs, symptoms, quality of life) that predict H^2^/R. Participants in the last 7 days of life were further assessed with BIA to assess hydration changes, and its relationship with clinical outcomes.

**Results:** 125 people participated (males n=74 (59.2%), females, n=51 (40.8%). BIVA demonstrated that baseline hydration status was normal in 58 (46.4%), ‘more-hydrated’ in 52 (41.6%) and ‘less hydrated’ in 13 (10.4%). Regression analysis demonstrated that less hydration (lower H^2^/R) was associated with female sex (Beta = -0.371, p<0.001), increased anxiety (Beta = - 0.135, <0.001), increased severity of physical signs (dry mouth, dry axilla, sunken eyes - Beta = -0.204, p<0.001), and increased breathlessness (Beta = -0.180, p<0.014). ‘More hydration’ (higher H^2^/R) was associated with oedema (Beta= 0.514, p<0.001) and increased pain (Beta = 0.156, p=0.039). Eighteen participants (14.4%) were in the last week of life. For dying participants, hydration status (H^2^/R) was not significantly different compared to baseline (n= 18, M= 49.55, SD= 16.00 vs. M= 50.96, SD= 12.13; t(17)= 0.636, p = 0.53) and was not significantly associated with agitation (r_s_ = -0.847, p = 0.740), pain (r_s_ = 0.306, p = 0.232) or respiratory tract secretions (r_s_ = -0.338, p = 0.185).

**Conclusions:** In advanced cancer, hydration status was associated with specific physical signs and symptoms. No significant associations between survival and hydration status were recorded. In the dying phase, hydration status did not significantly change compared to baseline, and was not associated with symptoms. Further work can use BIA/BIVA to standardise the process to identify clinically relevant outcomes for hydration studies, to establish a core outcome set to evaluate how hydration affects symptoms and quality of life in cancer.

**Key message:** We used bioelectrical impedance analysis (a non-invasive body composition assessment tool) to evaluate associations between hydration status and clinical outcomes in people with cancer. Hydration status was significantly associated with biological sex, physical signs, symptoms and psychological outcomes. In the dying phase, hydration status did not significantly change compared to baseline, and hydration status was not significantly associated with survival. The development of a standardised core outcome set for cancer hydration studies, to evaluate how hydration affects symptoms, quality of life and outcomes in cancer patients, will help to establish a meaningful evidence base for clinical practice.

## Introduction

Oral fluid intake commonly reduces in people with advanced cancer as they enter the dying phase of illness.[1] There is limited data describing how hydration status of advanced cancer is related to clinical outcomes[2] and inadequate evidence on the role of artificial hydration in clinical management.[3]

Bioelectrical impedance analysis (BIA) is a non-invasive body composition tool, which has usefulness in the assessment of hydration status, and its effects, in advanced cancer. [4, 5, 6, 7] BIA measures resistance to the flow of electrical current passed through the human body,[8] measuring resistance (R - the restriction to the flow of electrical current through the body, primarily related to the amount of water present in tissue) and reactance (Xc - resistive effect produced by the tissue interfaces and cell membranes). The impedance index (calculated from the equation = Height - H (m)^2^/R (Ohms)) is the best single predictor of total body water (TBW).[9, 10, 11, 12, 13, 14, 15, 16, 17, 18, 19, 20]

The BIA vector analysis (BIVA) RXc graph method involves standardization of BIA measurements by height, which are plotted as bivariate vectors with their confidence intervals, represented as ellipses on the R-Xc plane (Figure 1 - *The RXc graph with 95%, 75% and 50% tolerance ellipses).*The advantage of this method is that it allows for information to be obtained simultaneously about changes in tissue hydration or soft-tissue mass, independent of regression equations, or body weight. BIVA has been used to study hydration in a variety of different diseases[21, 22, 23, 24, 25, 26, 27, 28, 29] and to undertake general body composition assessments in people with lung cancer[28, 30] and cancers of the head and neck.[31]

**Figure 1.**
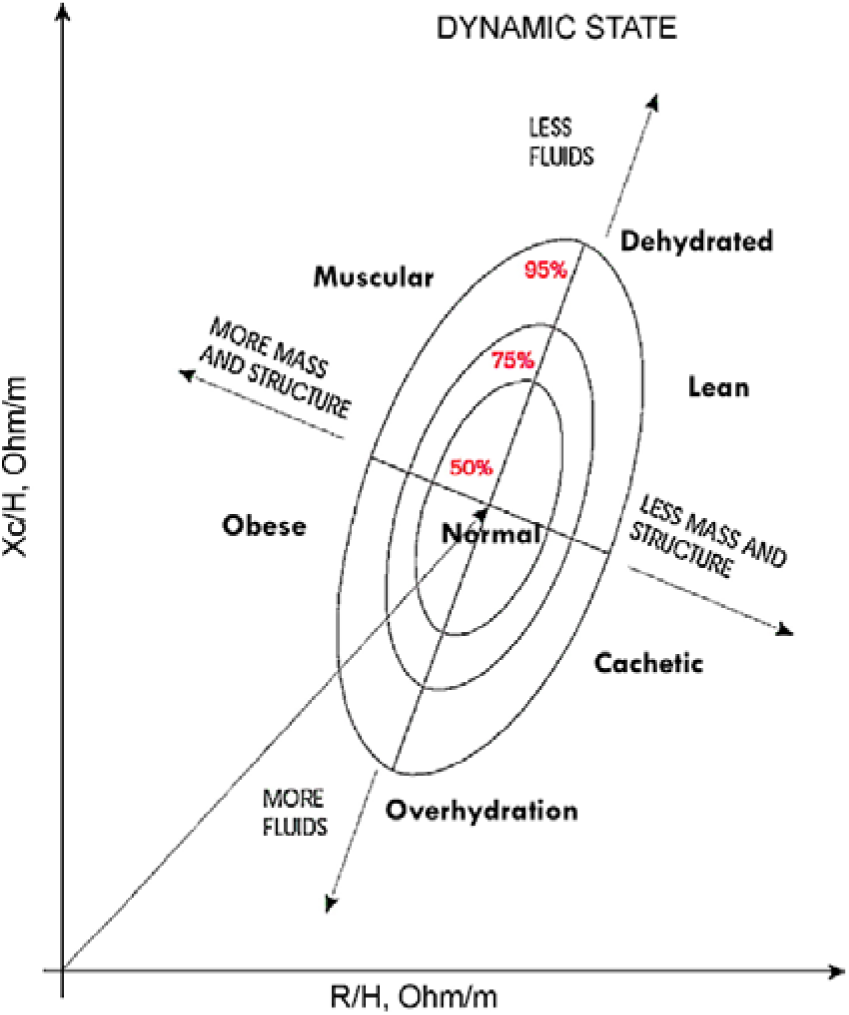
The RXc graph with 95%, 75% and 50% tolerance ellipses. Reproduced with permission).[34].

Our previous study used BIA/BIVA to report statistically significant associations between hydration status and clinical outcomes in advanced cancer.[5] Our previous study reported that lower hydration was associated with increased symptom intensity (dry mouth, thirst, taste and fatigue), increased physical signs (mouth moisture, sunken eyes and axilla dryness) and shorter survival (for people expected to live for ≥30 days), whereas higher hydration was associated with oedema.[5] However, our previous study did not evaluate potential associations between hydration and factors such as, myoclonus, quality of life or those lacking capacity. Consequently, our previous study was unable to describe how hydration status effected clinical outcomes in the dying (i.e., ≤7 days of life) as these individuals are commonly unable to provide consent, and hence, participate. Improving the evidence base to explain how hydration status affects several clinical outcomes in advanced cancer can potentially support healthcare professionals’ decision-making, and evaluate hydration interventions, particularly in the dying phase.[32] [33]

## Aim

The primary aim of this study is to evaluate hydration status, and its associations with clinical outcomes in advanced cancer. The secondary aim to assess hydration the effect of hydration on clinical outcomes in the dying phase (i.e., ≤7 days of life).

## Materials and Methods

We recruited participants from three UK specialist palliative care units (Academic Palliative Care Unit, Royal Liverpool University Hospital, Liverpool; Marie Curie Hospice West Midlands, Solihull, and Marie Curie Hospice Liverpool, Liverpool) between July 2017 and December 2020.

### Participants and eligibility

Eligibility criteria were: admission to specialist palliative care inpatient unit; age ≥18 years; cancer (proven by histology or radiology); palliative (i.e., no further curative treatment possible); and able to understand and communicate in English. Exclusion criteria were: implantable defibrillator devices; unable to provide fully informed consent; active transmissible infections; amputations; local wound infection or poor wound skin healing; and current antineoplastic treatment.

### Baseline assessments

We conducted assessments between 9am – 12pm. The following information was collected: age (years), biological sex, ethnicity (National Health Service England Ethnicity data categories[35]); primary site of cancer (according to the International Classification of Diseases[36]); and presence of metastatic disease. Participants received the following assessments:

### Bioelectrical impedance

BIA was conducted at the bedside using the AKERN BIA 101 Bio-impedance analyser. The BIA method involved a tetra-polar technique to deliver a single frequency electrical current of 50kHz (±5%). We conducted the testing procedure with methods described by Lukaski[37] and others.[38, 39] During testing, participants were lightly clothed, lying supine, without shoes or socks. We positioned their arms at a 30 degree angle from their body with their legs positioned 45 degrees away from each other. Two disposable pre-gelled aluminium electrodes were attached to the dorsum of their right hand (one placed on the edge of an imaginary line bisecting the ulnar head and the other on the middle finger proximal to the metacarpal-phalangeal) and two electrodes were placed to the dorsum of the right foot (one placed medially, to an imaginary line bisecting the medical malleolus at the ankle and the other proximal to the metatarsal-phalangeal joints). We calibrated the analyser daily using an impedance calibration circuit (R = 470 Ω, Xc = 90 Ω). BIA measurements of R, Xc, Phase Angle (PA) and the impedance index (calculated from the equation = Height - H (m)^2^/R (Ohms) were recorded.

### Dehydration Symptom Questionnaire (Appendix: Dehydration Symptom Questionnaire)

Participants reported dehydration-related symptom severity using a questionnaire developed by Burge,[40, 41] consisting of a numerical rating scale (0 least severe – 10 most severe) of four questions (thirst, dry mouth, unpleasant taste and fatigue) to measure symptom severity over the previous 24-hours.

### The Edmonton Symptom Assessment System (Appendix: The Edmonton Symptom Assessment System)

Participants rated the intensity of nine physical symptoms (pain, tiredness, drowsiness, nausea, lack of appetite, breathlessness, depression, anxiety, wellbeing) using a numerical rating (0 least severe – 10 most severe).[42]

### Morita Dehydration Assessment Score (Appendix: Morita Dehydration Assessment Score)

Dehydration severity was assessed by clinical examination (conducted by the researcher) using the approach described by Morita et al,[43] to assess moisture of mucous membranes (0: moist, 1: somewhat dry, 2: dry), axillary moisture (0: moist, 1: dry), and sunken eyes (0: normal, 1: slightly sunken, 2: sunken).[43] These signs have significant correlations with biological dehydration,[41, 44, 45, 46] with higher scores (range 0-5) indicating an increased risk of dehydration (previous studies have used a Morita cut-off of 2 to define an increased risk of dehydration[47, 48]).

### Peripheral oedema assessment (Appendix: Peripheral oedema assessment)

Oedema was assessed by observation (conducted by the researcher), recording oedema presence in the upper limb, lower limb, torso and/or abdomen (0 = none; 1 = present).

### Height

Height was measured, without shoes, to the nearest 0.1cm using a portable stadiometer (SECA 213 Height Measure / Stadiometer); we measured length in those unable to stand.

### Quality of Life assessment (Appendix: Quality of Life assessment)

Participants completed five areas of the Functional Assessment of Chronic Illness Therapy– Fatigue (FACIT - appendix). The areas evaluated were: ‘Lack of energy’; ‘I am worried my condition will worsen’; I am sleeping well’; ‘I am able to enjoy life’; and ‘I am content with the quality of my life right now’. Participants rated the intensity of fatigue and its related symptoms on a scale of 0 to 4 (0 = “not at all,” 4 = “very much”).[49]

### Performance status (Appendix: Performance status)

We used the Eastern Cooperative Oncology Group (ECOG - appendix) scale to describe the physical function of participants (0= fully active, 5 = dead).[50]

### Myoclonus assessment (Appendix: Myoclonus assessment)

We used section 2 of the Unified Myoclonus Rating Scale (UMRS – appendix) to determine the presence or absence of myoclonus at rest during a 10-second observation period.[51]

### Dying phase assessments

We defined ‘dying’ as an expected prognosis of ≤7 days. We used an advance consent process to enable participates to provide prior consent for research assessments in the dying phase. During the consent process, the participant chose a consultee (e.g., family caregiver, healthcare professional) who would provide assent to facilitate ongoing study participation if the participant was deemed (by their responsible clinical team) to be dying (e.g., identified by use of care plan to support management for the dying phase [52, 53]), and the participant was no longer able to provide consent for ongoing research participation. The following assessments were conducted in participants receiving ‘dying phase’ assessments: (1) a further BIA assessment and (2) a proxy measure of their comfort recorded by a healthcare professional or caregiver (Appendix: Proxy measurement of comfort recorded by a healthcare professional or caregiver).

### Statistical analysis

Statistical Package for the Social Sciences (SPSS) version 28.0 was used for standard calculations. Distributions of all variables were assessed for normality using the Shapiro-Wilk test (Appendix: Normality test). Parametric and non-parametric tests were used as appropriate to the data. Frequency analysis was conducted to compare differences between groups and variables using the chi-squared test, Student t-test and the Mann-Whitney U test. We conducted an exploratory univariate analysis of all data to explore associations with hydration (H^2^/R) with these variables (Appendix: Univariate test). A regression analysis was subsequently conducted to explore the influence of potential predictors on the H^2^/R.

### BIVA point graph analysis

For BIVA, the impedance vector (Z) was plotted as a bivariate vector from its components, R (X-axis) and Xc (Y-axis), after being standardized by height (H); this forms two correlated normal random variables (i.e. a bivariate Gaussian vector).[5, 54, 55] Elliptical probability regions of the mean vector are plotted on the RXc plane forming elliptical probability regions on the RXc plane, which are tolerance ellipses for individual vectors and confidence ellipses for mean vectors.[5, 8, 55, 56, 57, 58] Tolerance ellipses are the bivariate reference intervals of a normal population for an observation. The RXc graph features three tolerance ellipses: the median, the third quartile, and the 95th percentile (i.e., 50%, 75% and 95% of individual points). Participant data were plotted on the RXc point graph using the 50%, 75% and 95% tolerance ellipses from a non-cancer reference population.[8]). Hydration status can be described by dividing the BIA RXc normogram into three parallel sections, which correspond with the boundaries of each tolerance ellipse (*Figure 2-Classification of hydration status using the RXc graph and the 50th and 75th percentile tolerance ellipses*).[59] Individuals with vectors above the 50% tolerance ellipse were ‘less-hydrated’, participants with vectors in the central 50^th^ percentile ellipse were ‘normally-hydrated’, participants with vectors below the 50% tolerance ellipse were ‘more hydrated’.

**Figure 2.**
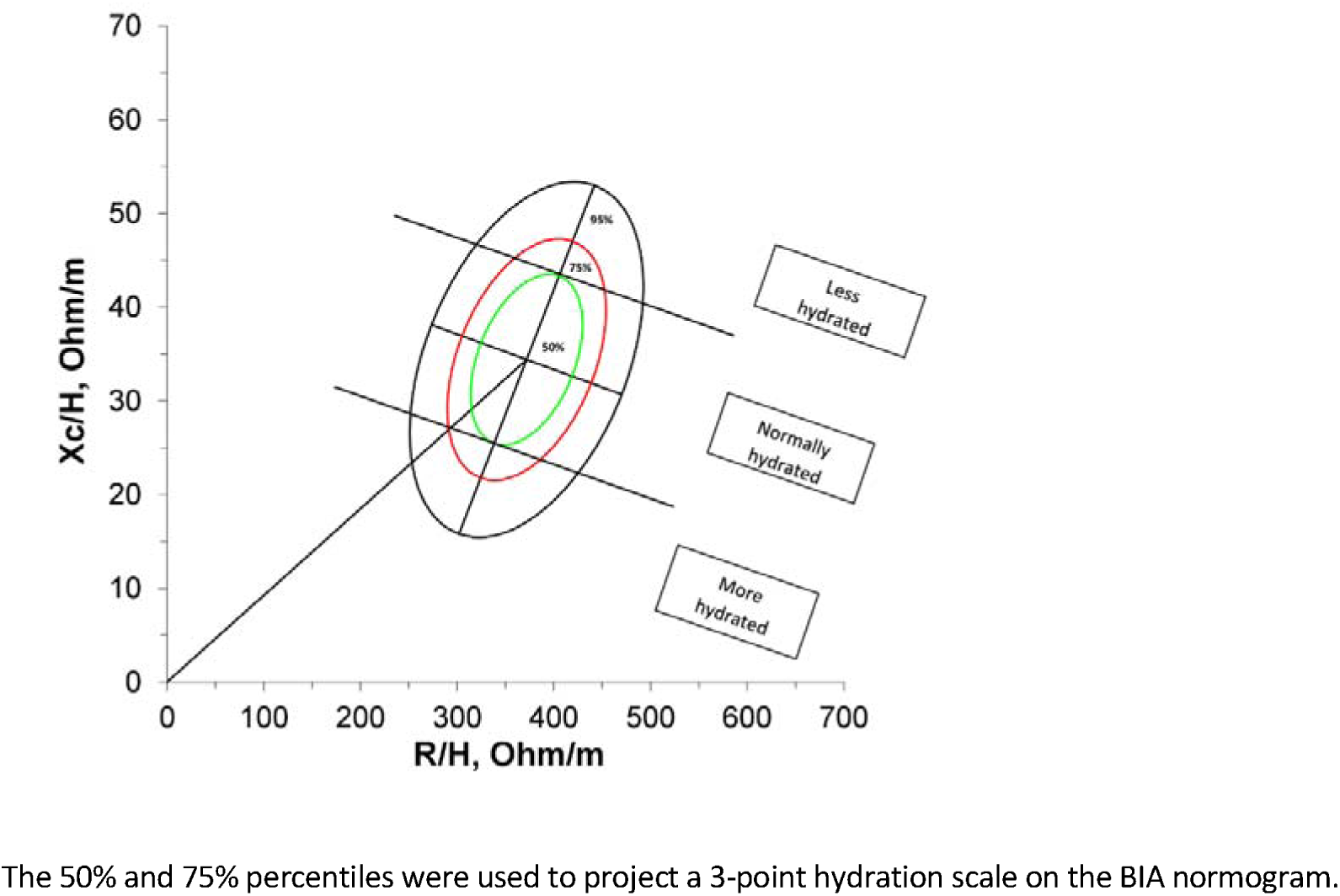
Classification of hydration status using the RXc graph and the 50th and 75th percentile tolerance ellipses.

### Backward regression analysis

A backward stepwise linear regression was used to explore the influence of potential predictors on the impedance index (used as a proxy for hydration). At each step variables were chosen based on p-values, and a p-value threshold of 0.1 was used to set a limit on the total number of variables included in the final model. We preselected nine variables, with evidence of association with hydration status in advanced cancer, for inclusion in the regression model. The selected variables were: age,[60, 61] sex,[62] anxiety,[63] Morita Dehydration score,[43] oedema,[5, 48, 64] thirst,[5] nausea,[1] pain[41] and breathlessness.[65] We included highly correlated variables from the univariate analysis to the pre-selected variables (to a maximum of one variable to 10 participants for the regression equation). The significance level was <0.05. BIVA statistical software for the analysis, developed by Professor Antonio Piccoli, University of Padova,[66] was used for the analysis.

### Analysis of dying participants receiving repeat assessments

Paired t-test analysis was used to compare change in hydration status from baseline to the dying phase assessments, by comparing the mean difference between the impedance index (H^2^/R). One sample Hotelling’s T^2^ test was used to determine statistical difference between baseline and dying phase assessments on the RXc graph. Spearman rank correlation coefficient was used to explore associations between hydration status and proxy symptom scores in the dying phase.

### Survival analysis

Survival was evaluated from baseline assessment date to death. All patients were followed up for 18 months following completion of the study. Kaplan-Meier analysis was used to analyse survival, according to the hydration status. The Cox proportional hazards model was used to assess the effect of hydration (H^2^/R) on survival, with adjustment for sex, age, baseline ECOG performance status, the presence of metastatic disease and cancer type.

### Sample size

An exploratory sample of 150 participants was selected to achieve a minimum of 10 participants for each item in the regression model (based on recommendations for observational studies).[67, 68] We aimed to recruit 20% of participants in the dying phase (i.e. last week of life) of their illness; therefore, 30 assessments were anticipated from a sample of 150.

## Results

### Demographics and baseline data

One hundred and twenty-five people participated (males n=74 (59.2%), females, n=51 (40.8%) (Figure 3 – recruitment flowchart). Primary cancer type was most commonly gastrointestinal (n=51/125, 50.8%), genitourinary (n=26, 20.8%) or lung (n=24, 19.2%). Most participants were white (n=119, 95.2%), with metastatic disease (n=78, 62.4%) and an ECOG performance status of 3 (n=68, 54.4%). Oedema was present in less than half of the participants (n=54, 43.2%) and a third had myoclonus (n=42, 33.6%) (*Table 1 - Demographic details of study participants*). Baseline assessment data are presented in Table 2. Dying phase assessments were conducted in 18 (14.4%) of participants (*Table 2 - Table 2. Baseline clinical assessment data*).

**Figure 3.**
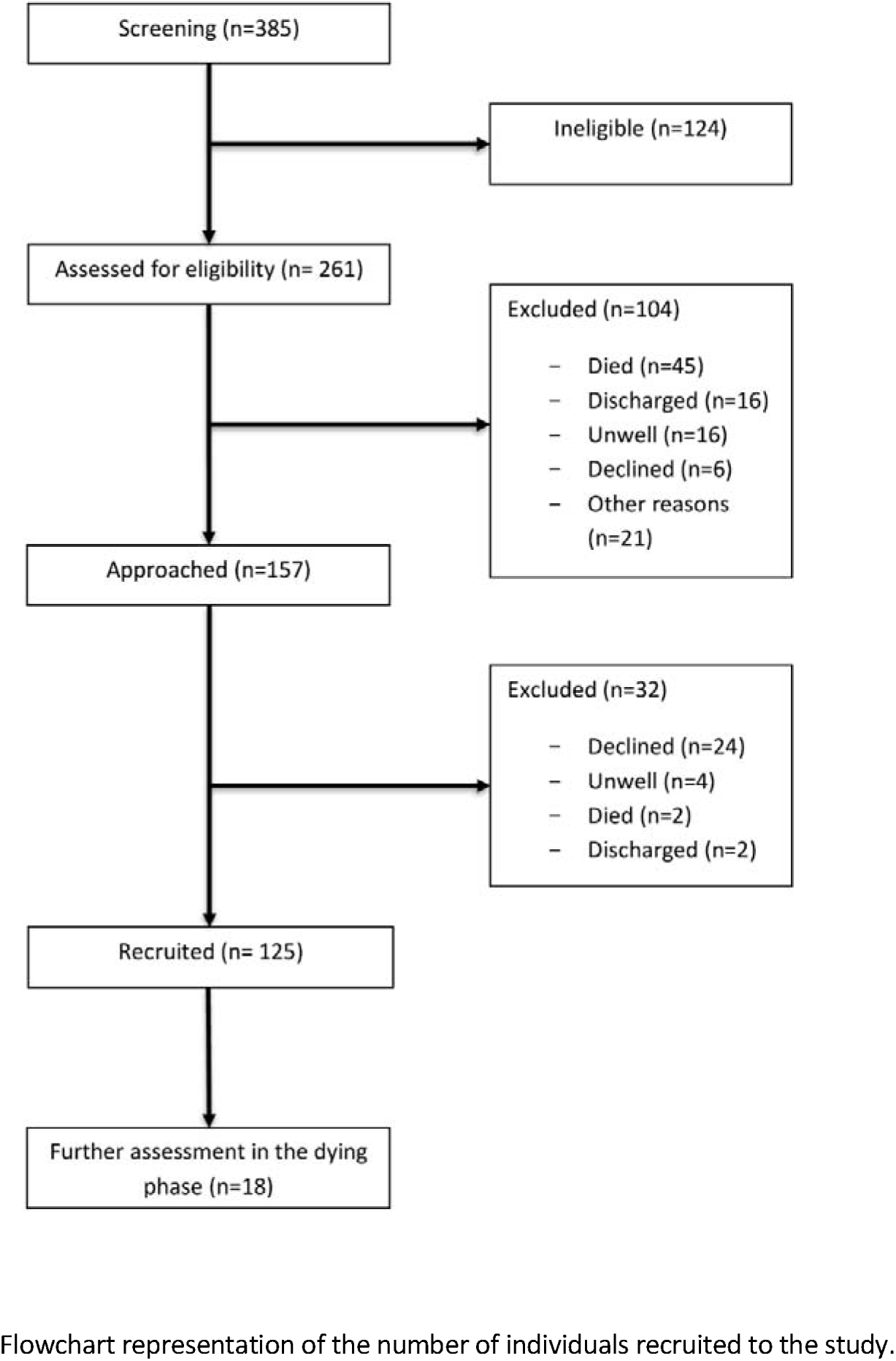
Recruitment flowchart.

**Table 1.**
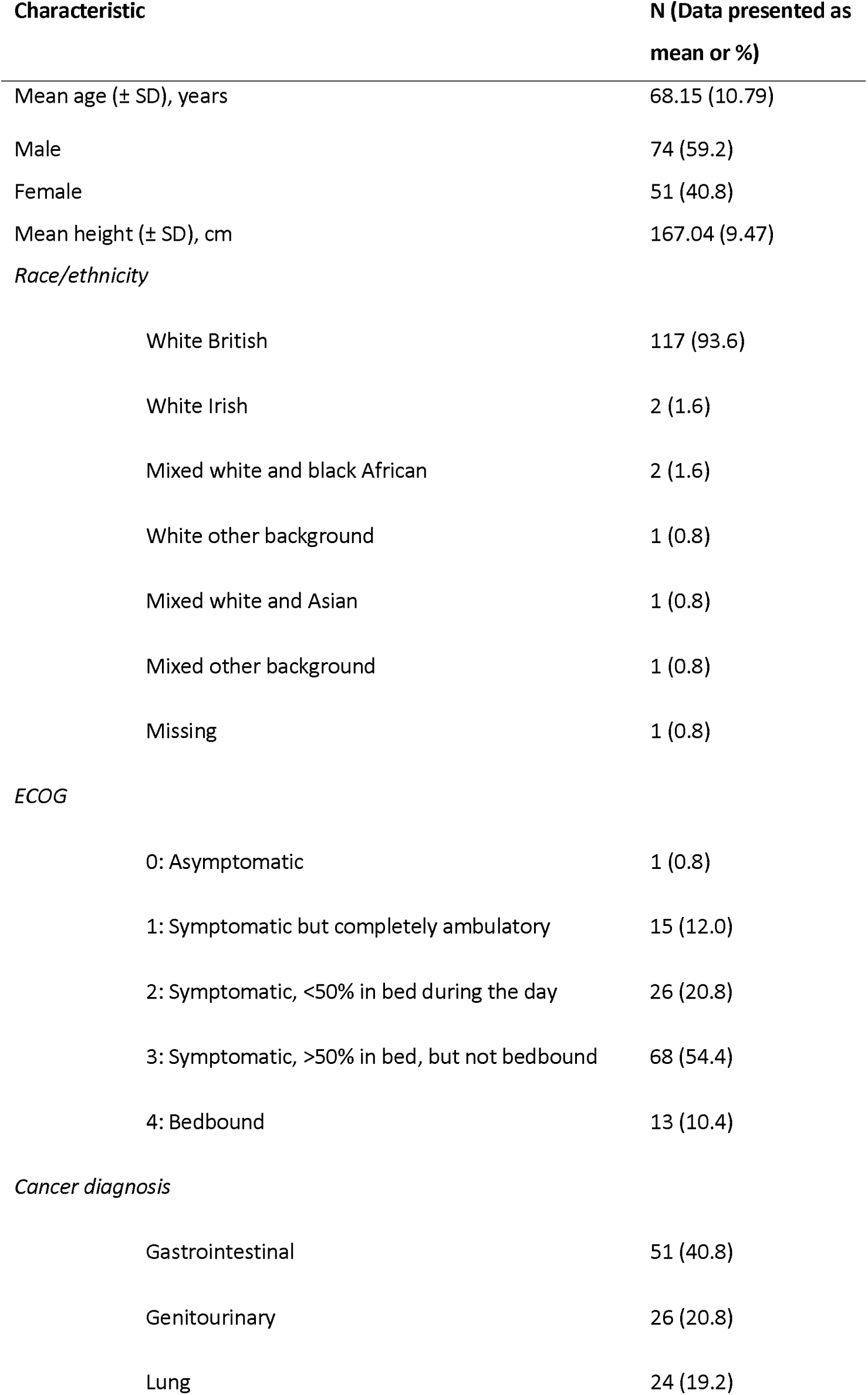
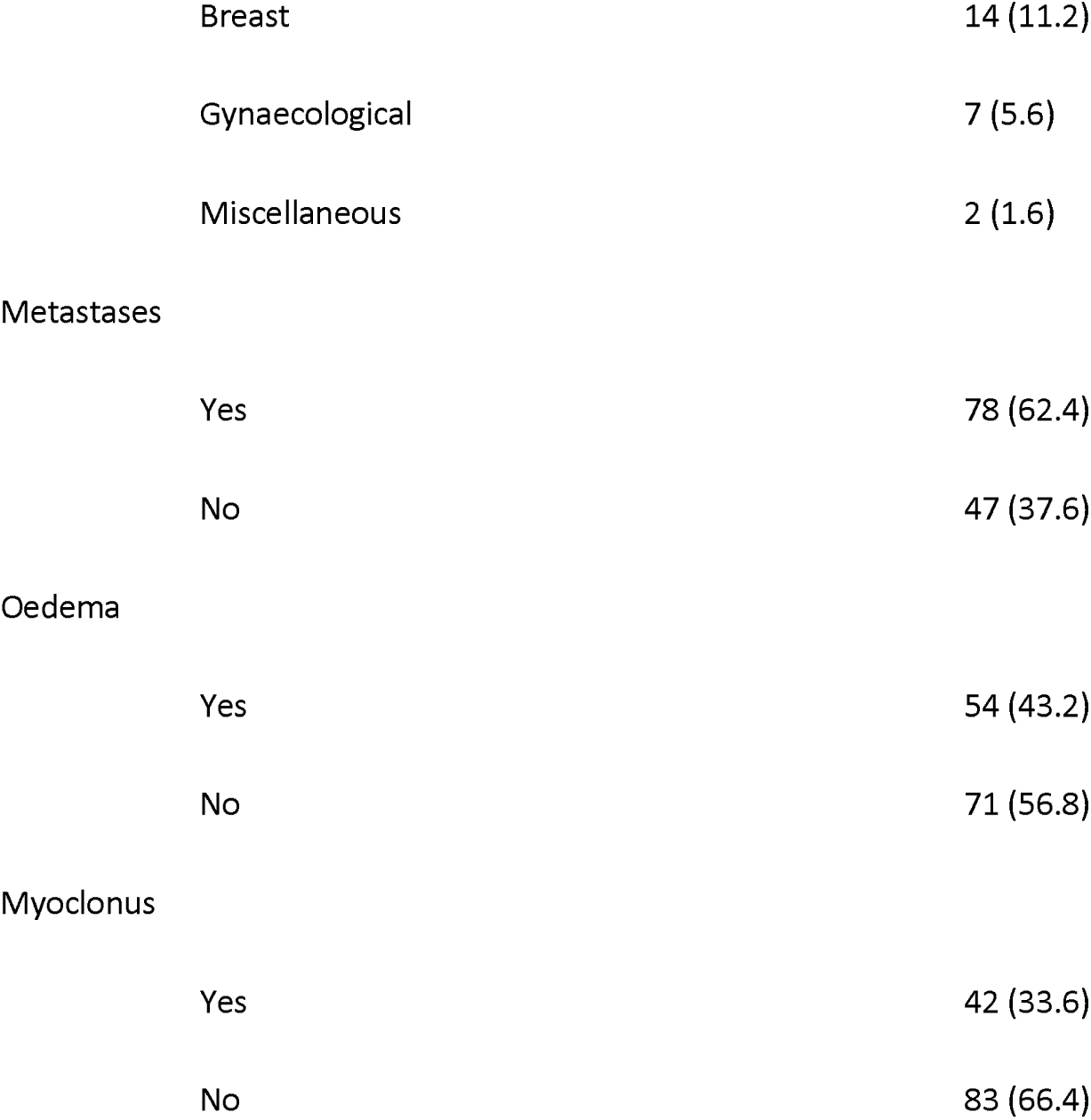
Demographic details of study participants.

| Characteristic | N (Data presented as mean or %) |
| --- | --- |
| Mean age (± SD), years | 68.15 (10.79) |
| Male | 74 (59.2) |
| Female | 51 (40.8) |
| Mean height (± SD), cm | 167.04 (9.47) |
| *Race/ethnicity* |  |
| White British | 117 (93.6) |
| White Irish | 2 (1.6) |
| Mixed white and black African | 2 (1.6) |
| White other background | 1 (0.8) |
| Mixed white and Asian | 1 (0.8) |
| Mixed other background | 1 (0.8) |
| Missing | 1 (0.8) |
| *ECOG* |  |
| 0: Asymptomatic | 1 (0.8) |
| 1: Symptomatic but completely ambulatory | 15 (12.0) |
| 2: Symptomatic, <50% in bed during the day | 26 (20.8) |
| 3: Symptomatic, >50% in bed, but not bedbound | 68 (54.4) |
| 4: Bedbound | 13 (10.4) |
| *Cancer diagnosis* |  |
| Gastrointestinal | 51 (40.8) |
| Genitourinary | 26 (20.8) |
| Lung | 24 (19.2) |
| Breast | 14 (11.2) |
| Gynaecological | 7 (5.6) |
| Miscellaneous | 2 (1.6) |
| Metastases |  |
| Yes | 78 (62.4) |
| No | 47 (37.6) |
| Oedema |  |
| Yes | 54 (43.2) |
| No | 71 (56.8) |
| Myoclonus |  |
| Yes | 42 (33.6) |
| No | 83 (66.4) |

**Table 2.**
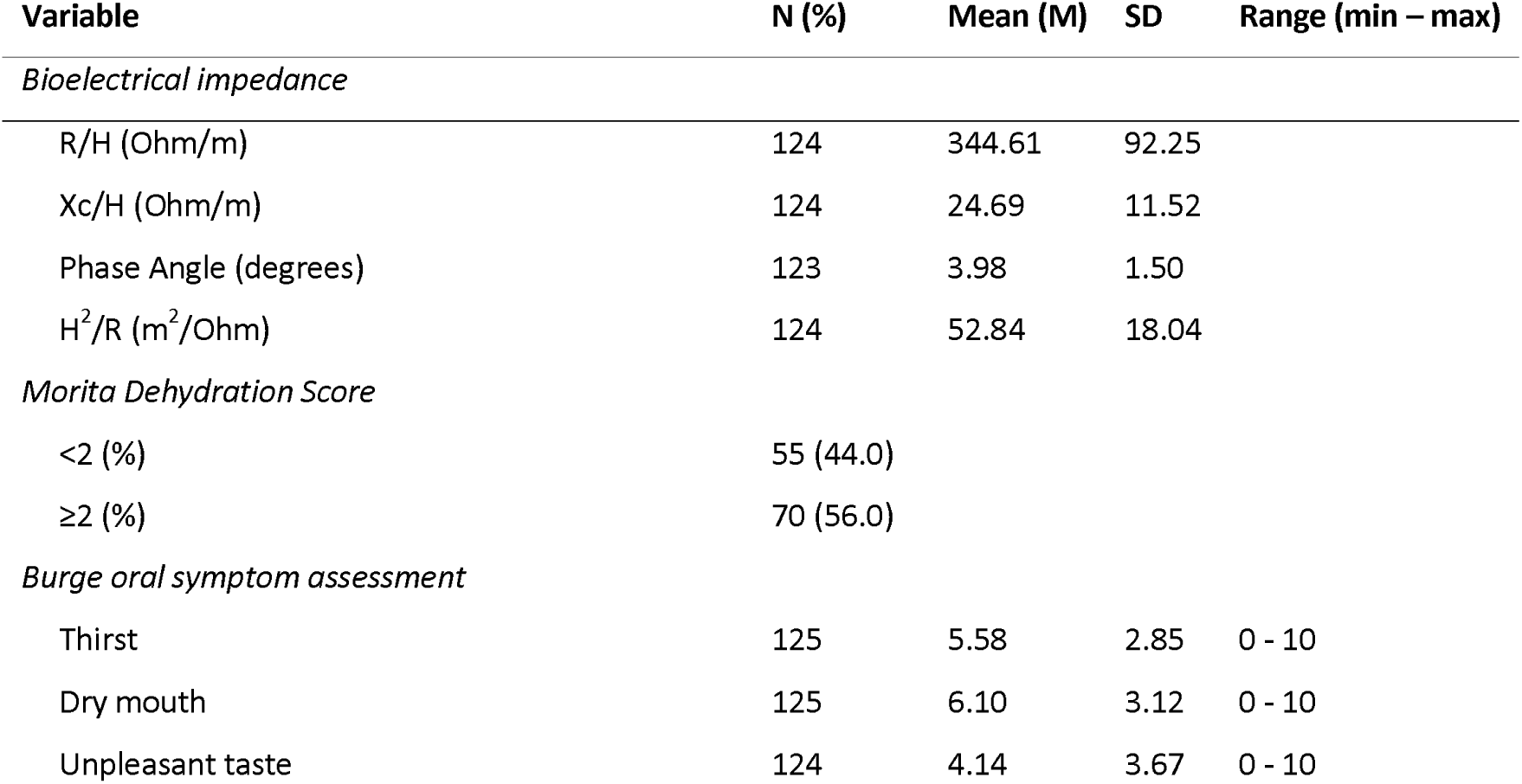
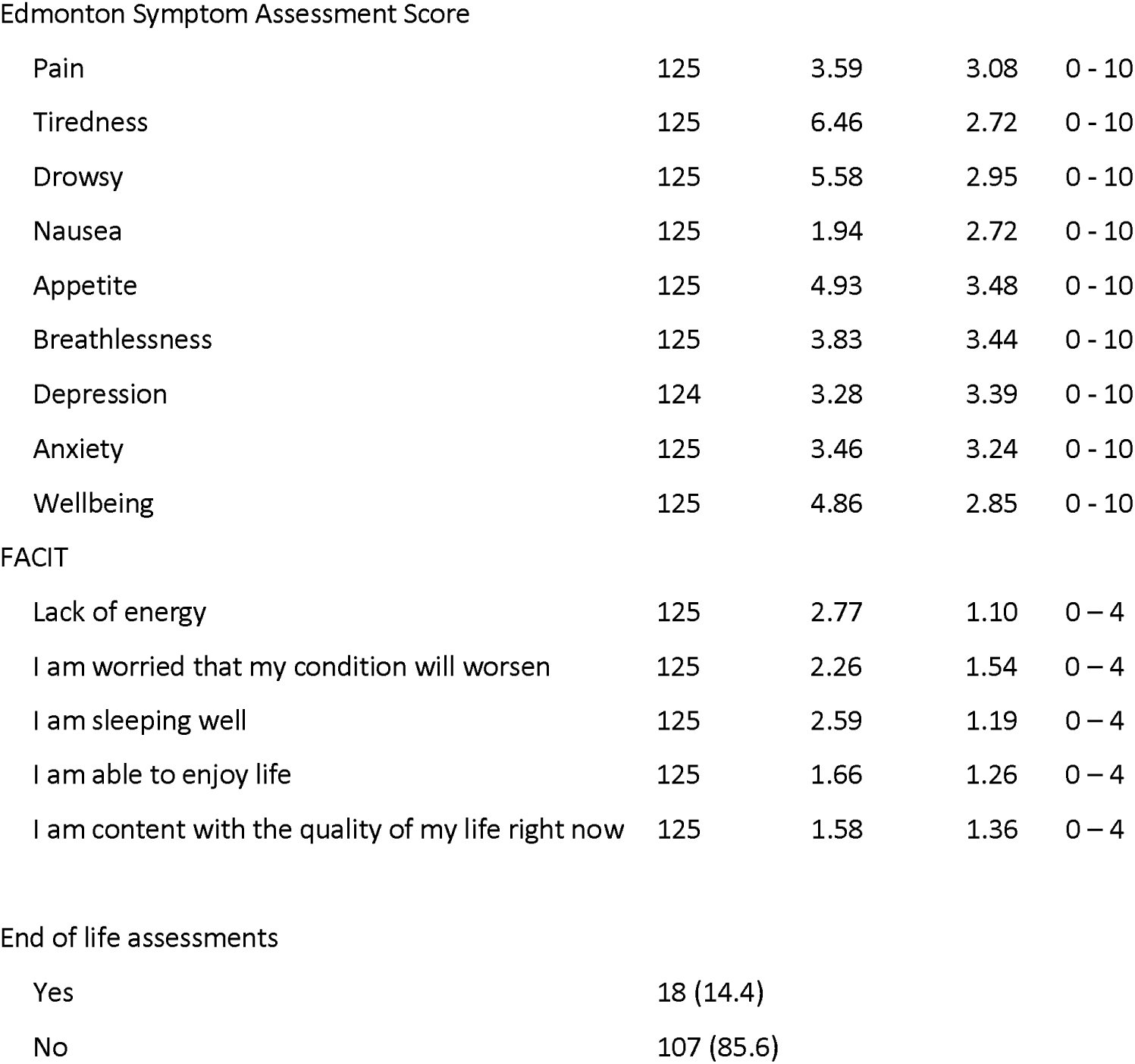
Baseline clinical assessment data.

| Variable | N (%) | Mean (M) | SD | Range (min – max) |
| --- | --- | --- | --- | --- |
| *Bioelectrical impedance* |  |  |  |  |
| R/H (Ohm/m) | 124 | 344.61 | 92.25 |  |
| Xc/H (Ohm/m) | 124 | 24.69 | 11.52 |  |
| Phase Angle (degrees) | 123 | 3.98 | 1.50 |  |
| H<sup>2</sup>/R (m<sup>2</sup>/Ohm) | 124 | 52.84 | 18.04 |  |
| *Morita Dehydration Score* |  |  |  |  |
| <2 (%) | 55 (44.0) |  |  |  |
| ≥2 (%) | 70 (56.0) |  |  |  |
| *Burge oral symptom assessment* |  |  |  |  |
| Thirst | 125 | 5.58 | 2.85 | 0 - 10 |
| Dry mouth | 125 | 6.10 | 3.12 | 0 - 10 |
| Unpleasant taste | 124 | 4.14 | 3.67 | 0 - 10 |
| Edmonton Symptom Assessment Score |  |  |  |  |
| Pain | 125 | 3.59 | 3.08 | 0 - 10 |
| Tiredness | 125 | 6.46 | 2.72 | 0 - 10 |
| Drowsy | 125 | 5.58 | 2.95 | 0 - 10 |
| Nausea | 125 | 1.94 | 2.72 | 0 - 10 |
| Appetite | 125 | 4.93 | 3.48 | 0 - 10 |
| Breathlessness | 125 | 3.83 | 3.44 | 0 - 10 |
| Depression | 124 | 3.28 | 3.39 | 0 - 10 |
| Anxiety | 125 | 3.46 | 3.24 | 0 - 10 |
| Wellbeing | 125 | 4.86 | 2.85 | 0 - 10 |
| FACIT |  |  |  |  |
| Lack of energy | 125 | 2.77 | 1.10 | 0 - 4 |
| I am worried that my condition will worsen | 125 | 2.26 | 1.54 | 0 - 4 |
| I am sleeping well | 125 | 2.59 | 1.19 | 0 - 4 |
| I am able to enjoy life | 125 | 1.66 | 1.26 | 0 - 4 |
| I am content with the quality of my life right now | 125 | 1.58 | 1.36 | 0 - 4 |

End of life assessments
|  |  |
| --- | --- |
| Yes | 18 (14.4) |
| No | 107 (85.6) |

### Hydration assessment and BIVA

Hydration status was normal in 58 (46.4%), ‘more-hydrated’ in 52 (41.6%) and ‘less hydrated’ in 13 (10.4%) (*Table 3 - Classification of hydration as a three-item scale according to the RXc graph scale; Figure 3 - Bioimpedance vector positions for females on the RXc point graph (N = 51); Figure 4. Bioimpedance vector positions for males on the RXc point graph (N = 74))*. Bioimpedance data for participants (and the reference population used for the comparative analysis) are presented in Table 4 (Table 4 - Bioelectrical impedance data).

**Figure 3a.**
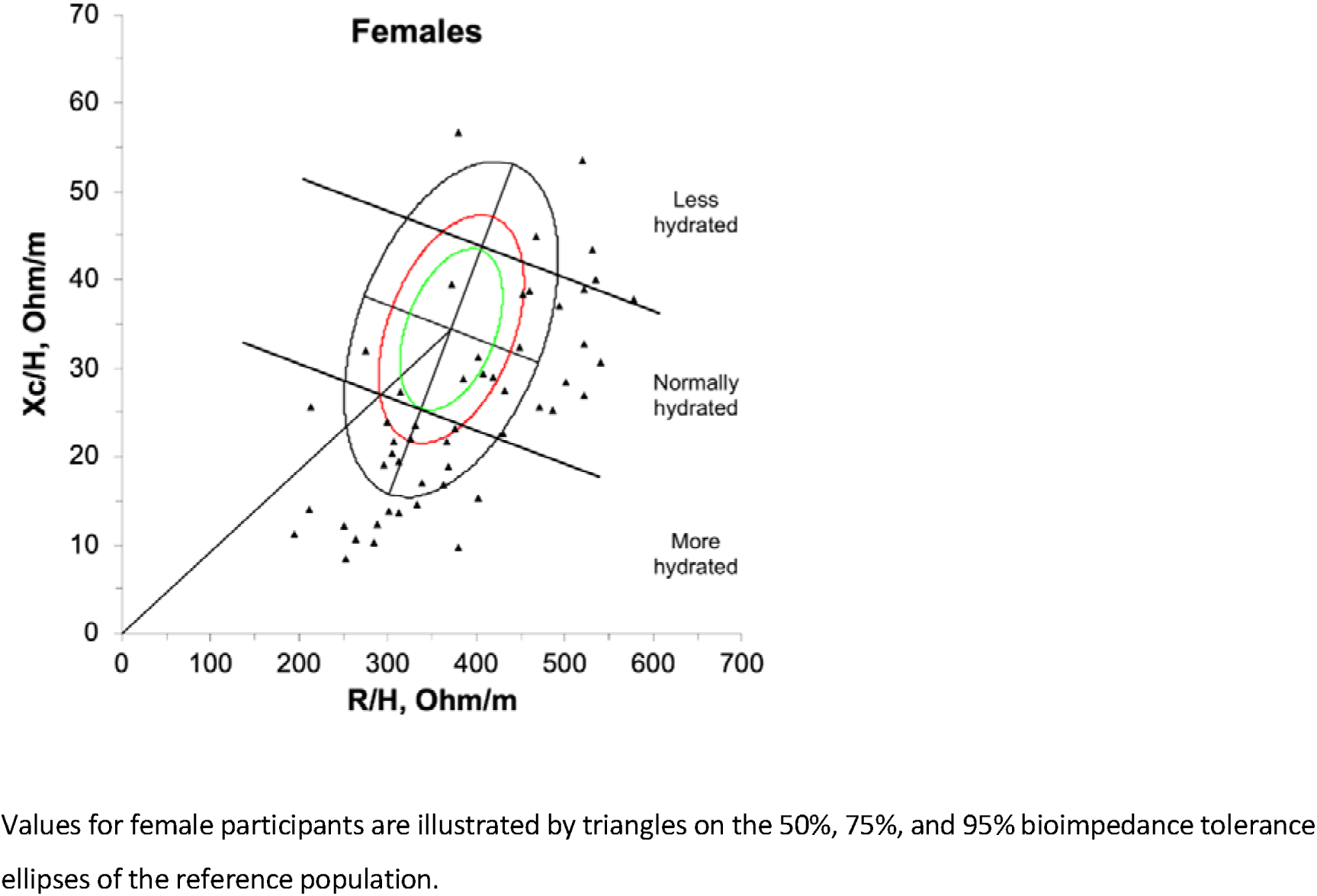
Bioimpedance vector positions for females on the RXc point graph (N = 51).

**Figure 4.**
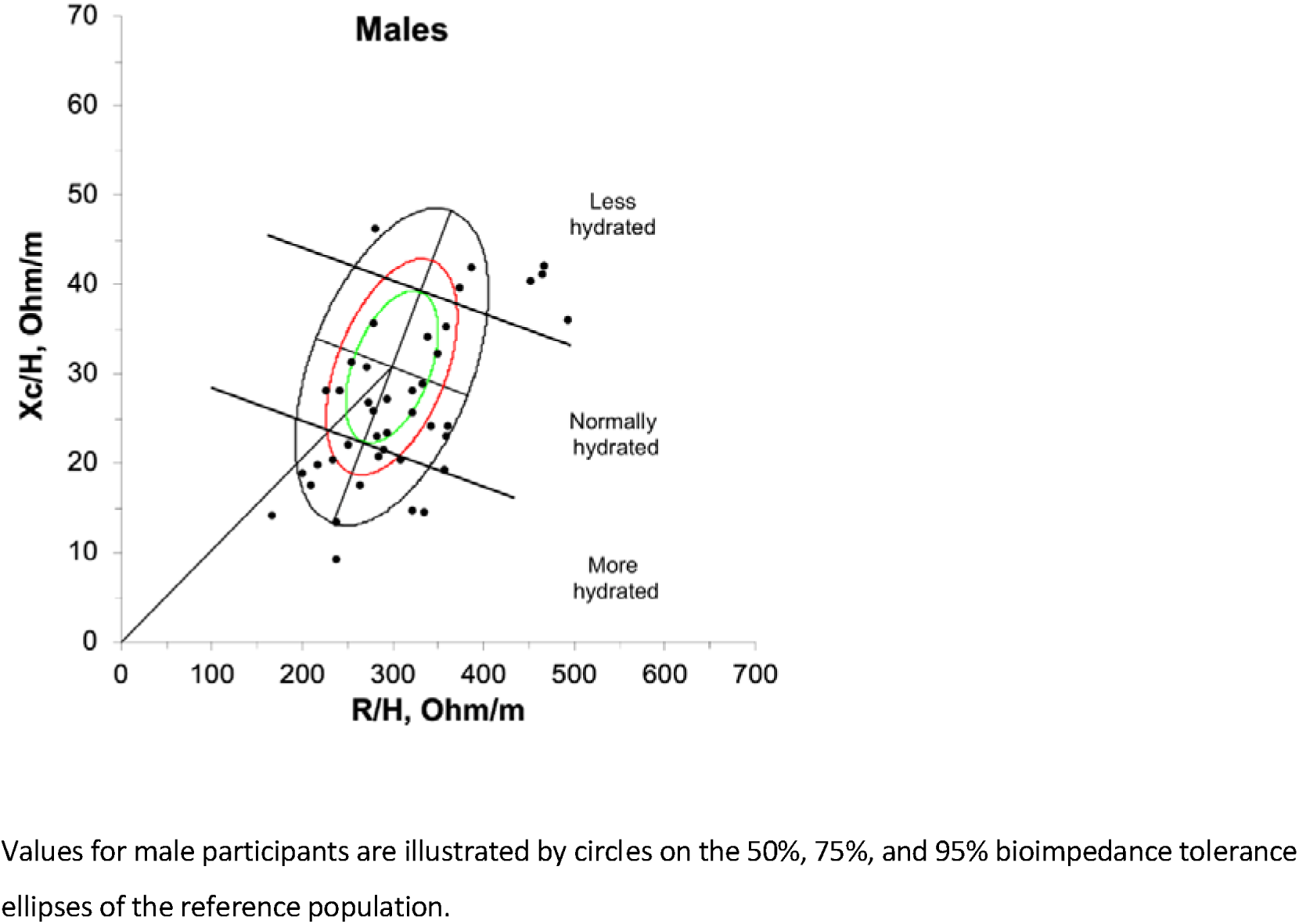
Bioimpedance vector positions for males on the RXc point graph (N = 74)

**Table 3.** Classification of hydration as a three-item scale according to the RXc graph scale.

| Hydration status | Male | Female | Total (%) |
| --- | --- | --- | --- |
| Less hydrated | 7 | 6 | 13 (10.4) |
| Normally hydrated | 38 | 20 | 58 (46.4) |
| More hydrated | 27 | 25 | 52 (41.6) |
| Missing | 2 | 0 | 2 (1.6) |
| Total | 72 | 51 | 125 |

**Table 4.** Bioelectrical impedance data.

| Sample data | R -Ohm (SD) | Xc – Ohm (SD) | Phase angle –<br>degrees (SD) | R/H - Ohm/m<br>(SD) | Xc/H - Ohm/m<br>(SD) | Impedance<br>index | Reference population data<sup>a</sup> |  |  |
| --- | --- | --- | --- | --- | --- | --- | --- | --- | --- |
|  |  |  |  |  |  |  | N | R/H - Ohm/m<br>(SD) | Xc/H - Ohm/m<br>(SD) |
| **Baseline data** |  |  |  |  |  |  |  |  |  |
| All patients (n = 125) | 572.02 (142.6) | 40.99 (18.76) | 3.98 (1.50) | 344.61 (92.25) | 24.70 (11.52) | 52.84 (18.04) | - | - | - |
| Males (n = 74) | 542.55 (131.44) | 40.68 (19.53) | 4.14 (1.62) | 317.55 (76.90) | 23.89 (11.66) | 58.02 (18.70) | 354 | 371.9 (43.2) | 30.8 (7.2) |
| Female (n = 51) | 614.20 (148.55) | 41.43 (17.79) | 3.75 (1.30) | 383.33 (99.10) | 25.85 (11.34) | 45.43 (14.20) | 372 | 298.6 (49.0) | 34.4 (7.7) |
| **Repeat data** |  |  |  |  |  |  |  |  |  |
| All patients (n =18) |  |  |  |  |  | 49.55 (16.00) | - | - | - |
| Males (n = 9) |  |  |  |  |  | 56.15 (18.58) | 354 | 371.9 (43.2) | 30.8 (7.2) |
| Female (n = 9) |  |  |  |  |  | 42.96 (10.05) | 372 | 298.6 (49.0) | 34.4 (7.7) |

<sup>a</sup> The Piccoli et al., 1995[8] reference population data was used for comparison with the outcomes from this study.

### Univariate analysis (Appendix - Univariate analysis)

The univariate analysis identified that lower H^2^/R (lower total body water) was associated with female sex (r_s_ -0.378, p<0.001); reduced appetite (r_s_ -0.273 p<0.002); increased anxiety (r_s_ -0.192, p<0.032); increased mouth dryness (r_s_ -0.242, p<0.007); higher Morita dehydration score (r_s_ -0.320, p<0.01); increased concern of participants that their condition will worsen over the next 7 days (r_s_ -0.19, p=0.034). Higher H^2^/R (suggesting higher total body water) was significantly associated with oedema (r_s_ 0.509, p<0.001) and improved sleep (r_s_ -0.235, p=0.009).

### Backward regression analysis

Three variables from the univariate analysis (reduced appetite, improved sleep and ‘worry my condition will worsen’) were added to the nine variables, preselected, for the backward regression analysis. Backward stepwise linear regression reduced 12 variables to 5, to identify a regression equation to predict H^2^/R (F2(6, 117) = 22.168, p <0.001), with an R^2^ of 0.532). This equation demonstrated that lower H^2^/R (less hydration) was associated with female sex (Beta = -0.371, p<0.001), more anxiety (Beta = -0.135, <0.001), more physical signs (dry mouth, dry axilla, sunken eyes - Beta = -0.204, p<0.001), and increased breathlessness (Beta = -0.180, p<0.014). Higher H^2^/R (more hydration) was associated with oedema (Beta= 0.514, p<0.001) and increased pain (Beta = 0.156, p=0.039) (Table 5 - Backward regression analysis of the impedance index (H^2^/R)).

**Table 5:**
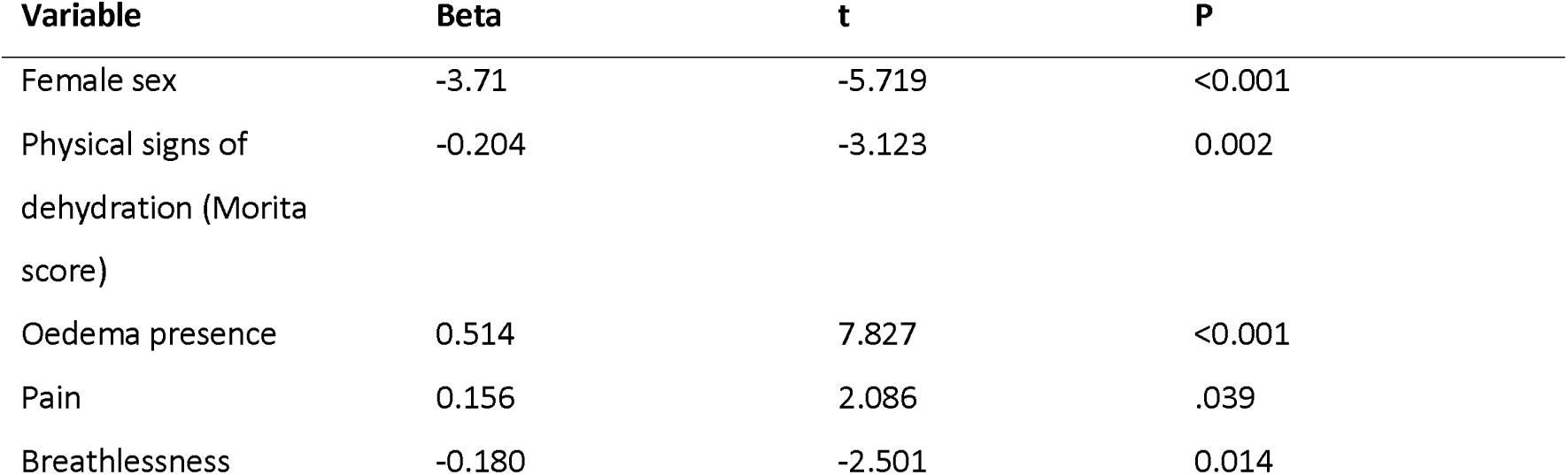
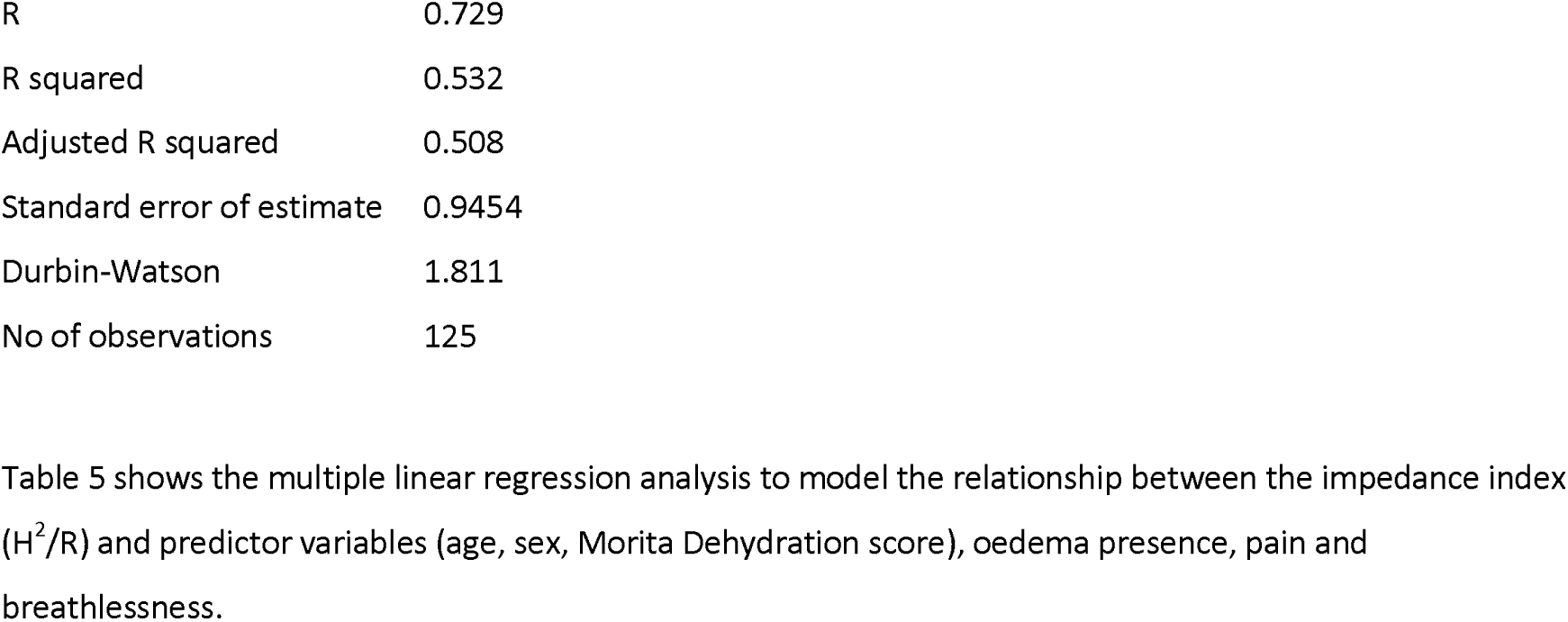
Backward regression analysis of the impedance index (H^2^/R)

| Variable | Beta | t | P |
| --- | --- | --- | --- |
| Female sex | -3.71 | -5.719 | <0.001 |
| Physical signs of dehydration (Morita score) | -0.204 | -3.123 | 0.002 |
| Oedema presence | 0.514 | 7.827 | <0.001 |
| Pain | 0.156 | 2.086 | .039 |
| Breathlessness | -0.180 | -2.501 | 0.014 |

|  |  |
| --- | --- |
| R | 0.729 |
| R squared | 0.532 |
| Adjusted R squared | 0.508 |
| Standard error of estimate | 0.9454 |
| Durbin-Watson | 1.811 |
| No of observations | 125 |

Table 5 shows the multiple linear regression analysis to model the relationship between the impedance index ( $H^2/R$ ) and predictor variables (age, sex, Morita Dehydration score), oedema presence, pain and breathlessness.

### Assessments in the dying phase

Eighteen (14.4%) dying participants received further assessment (Table 6 - Bioimpedance assessment data at the end of life). Paired t-test demonstrated that hydration status (H^2^/R) of dying participants illness was not significantly different to the baseline assessments of the participants (n= 18, M= 49.55, SD= 16.00 vs. M= 50.96, SD= 12.13; t(17)= 0.636, p = 0.53). Furthermore, the Hotelling’s test of paired BIVA vector data (comparing baseline and repeat hydration assessment) demonstrated no significant difference of hydration status, as the 95% confidence ellipse crossed the origin (i.e., 0,0 on the RXc graph) (Appendix: The Hotelling’s test of paired BIVA vector data to compare baseline and end of life care assessments).[69] The H^2^/R was not significantly associated with agitation (r_s_ = -0.847, p = 0.740), pain (r_s_ = 0.306, p = 0.232) and respiratory tract secretions (r_s_ = -0.338, p = 0.185) in dying participants, which suggests that that hydration status was not associated with these symptoms in the dying phase. (Table 7 - Spearman rank correlation coefficients to describe association between impedance index and symptoms in dying participants).

**Table 6:** Bioimpedance assessment data at the end of life.

| Variable | N | Mean<br>(M) | SD | Standard<br>error | Paired t-test |  |  |
| --- | --- | --- | --- | --- | --- | --- | --- |
|  |  |  |  |  | t | df | Sig (two-tailed) |
| Impedance index | 18 | 50.9631 | 12.13720 | 2.86077 | 0.636 | 17 | 0.533 |
| Follow-up<br>impedance index | 18 | 49.5539 | 16.00192 | 3.77169 |  |  |  |

**Table 7:** Spearman rank correlation coefficients to describe association between impedance index (H^2^/R) and symptoms in dying participants.

| Variable | N | Correlation coefficient<br>($r_s$) | Sig (two-tailed) |
| --- | --- | --- | --- |
| Agitation | 18 | -0.847 | 0.740 |
| Pain | 17 | 0.306 | 0.232 |
| Respiratory tract<br>secretions | 17 | -0.338 | 0.185 |

### Survival analysis

123 (98.4%) participants died by the end of the follow-up period. Median survival for the sample was 35 days (IQR 17 - 106) (Table 8 - Survival data for participants according to hydration status). Median survival was shortest in ‘more hydrated’ participants (32 days, IQR 15 - 72) and longest in those ‘less hydrated’ (45 days, IQR 21 - 117). No significant associations between survival and hydration status were recorded (*Figure 5. Kaplan-Meier graph showing survival time in days according to BIVA determined hydration status*). Multivariate Cox regression survival analysis did not demonstrate significant associations with survival with age, sex performance status, cancer type, metastatic disease, and impedance index (*Table 9 - Multivariate cox regression analysis for death according to age, sex performance status, cancer type, metastatic disease and impedance index)*.

**Figure 5.**
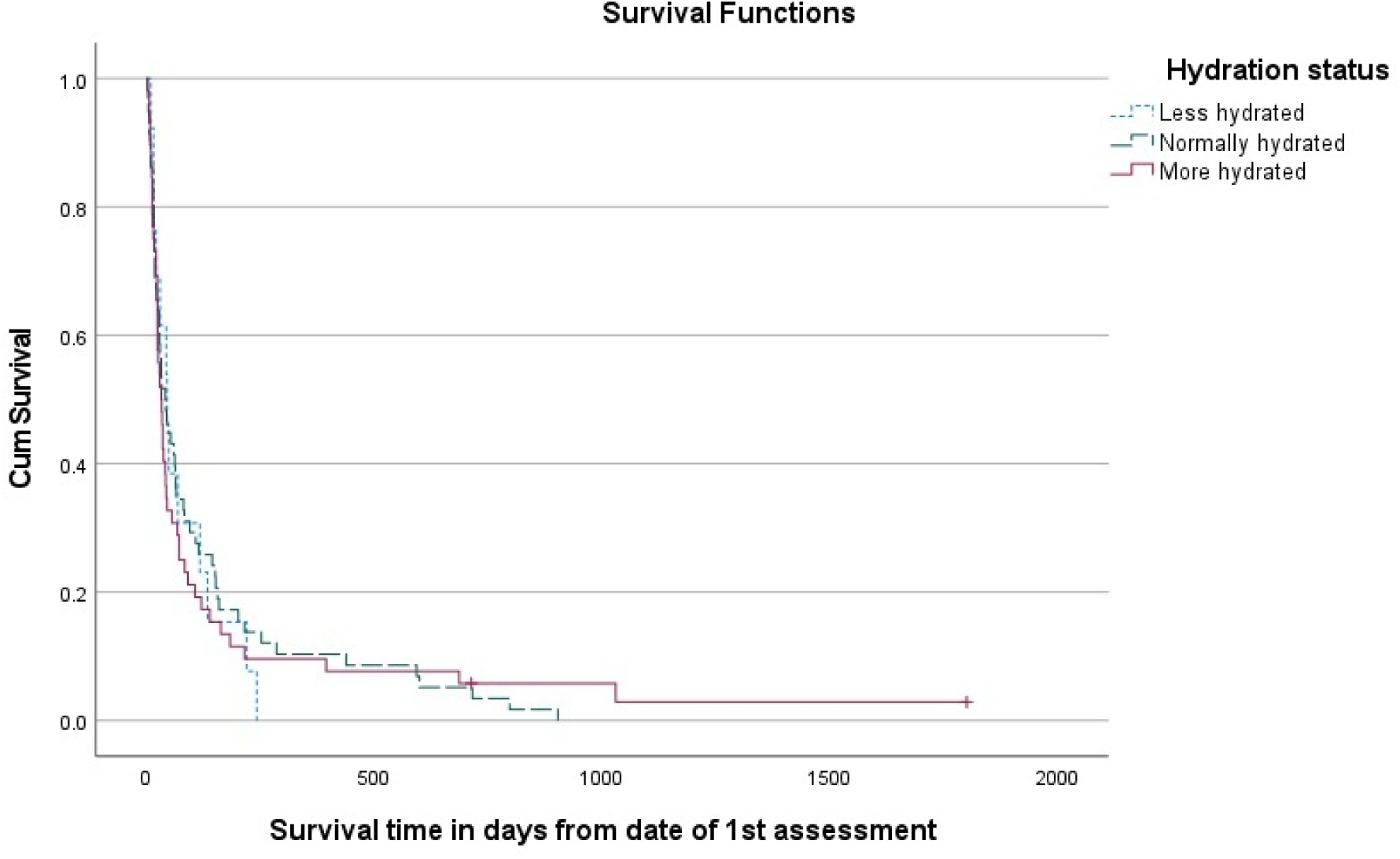
Kaplan-Meier graph showing survival time in days according to BIVA determined hydration status (χ^2^ = 0.17, P= 0.93)

**Table 8.** Univariate survival analysis of participants according to BIVA determined hydration status.

| Subgroup | N | Median survival in days (IQR) | Hazard ratio (95% CI) | p |
| --- | --- | --- | --- | --- |
| Overall | 123 | 35.0 (17.0 – 106) |  |  |
| *Hydration classification according to three BIVA classifications* |  |  |  |  |
| Less hydrated | 13 | 45.0 (21 – 117) | 1.00 (ref) | 0.94 |
| Normal | 58 | 41.0 (17 – 144) | 0.94 (0.51, 1.72) | 0.87 |
| More hydrated | 52 | 32.0 (15 – 72) | 1.01 (0.55, 1.87) | 0.98 |

**Table 9:** Multivariate cox regression analysis for death according to age, sex performance status, cancer type, metastatic disease and impedance index.

| Variable | Hazard Ratio (95% CI) | p |
| --- | --- | --- |
| Age (years) | 1.005 (0.99, 1.02) | 0.57 |
| Female | 1.03 (0.67, 1.58) | 0.90 |
| ECOG 1 | - | 0.11 |
| ECOG (2 vs. 1) | 0.96 (0.12, 7.73) | 0.97 |
| ECOG (3 vs. 1) | 1.95 (0.26, 14.78) | 0.88 |
| ECOG (4 vs. 1) | 1.54 (0.18, 13.30) | 0.52 |
| Cancer type (GI) | - | 0.83 |
| Cancer type (Gynae vs. GI) | 0.99 (0.99, 0.34) | 2.92 |
| Cancer type (Lung vs. GI) | 0.70 (1.11, 0.65) | 1.91 |
| Cancer type (GU vs. GI) | 0.64 (0.89, 0.54) | 1.46 |
| Cancer type (Breast vs GI) | 0.41 (0.75, 0.38) | 1.49 |
| Cancer type (Misc vs. GI) | 0.36 (0.45, 8.97) | 0.36 |
| Metastases present | 1.45 (0.97, 2.18) | 0.07 |
| H<sup>2</sup>/R (m<sup>2</sup>/Ohm) | 1.00 (0.99, 1.01) | 0.72 |

## Discussion

### Summary of findings from the entire cohort

Our findings highlight that baseline hydration status was normal in 58 (46.4%), ‘more-hydrated’ in 52 (41.6%) and ‘less hydrated’ in 13 (10.4%). Hydration status was associated with several clinical outcomes, with lower hydration status (lower H^2^/R) associated with female sex (Beta = -0.371, p<0.001), increased anxiety (Beta = -0.135, <0.001), increased severity of physical signs (dry mouth, dry axilla, sunken eyes - Beta = -0.204, p<0.001), and increased breathlessness (Beta = -0.180, p<0.014). ‘More hydration’ (higher H^2^/R) was associated with oedema (Beta= 0.514, p<0.001) and increased pain (Beta = 0.156, p=0.039). No significant associations between hydration status and survival were recorded.

### Summary of findings from the dying participants

Eighteen participants of our sample were classified as dying and received further assessment (n=18, 14.4%). Hydration status (H^2^/R) of dying participants was not significantly different compared to their baseline assessment (n= 18, M= 49.55, SD= 16.00 vs. M= 50.96, SD= 12.13; t(17)= 0.636, p = 0.53). The H^2^/R was not significantly associated with agitation (r_s_ = -0.847, p = 0.740), pain (r_s_ = 0.306, p = 0.232) and respiratory tract secretions (r_s_ = - 0.338, p = 0.185) in dying participants.

### New knowledge

This paper builds on our previous BIA/BIVA work which described significant associations between status and clinical outcomes in advanced cancer.[5] We provide bioimpedance data an adults with advanced cancer, for use as reference population data in future BIA/BIVA reference. Further, we demonstrate opportunities for palliative care research, through body composition assessment using BIA/BIVA, and the advance consent method to research the dying.

### Comparison with previous work

Consistent with previous work, hydration status was associated with physiological outcomes.[5] Females had comparatively less body water (lower H^2^/R) than men, which (Beta = -0.371, p<0.001) has physiological explanation, as women generally have less lean-muscle mass compared to men. Intracellular water is most present in lean muscle; therefore, women comparatively have less body water than men as they generally have less muscle mass.[8, 57, 70]

Lower H^2^/R (less hydration) was associated with more anxiety, a finding consistent with previous non-cancer research that describes how water consumption is associated with improved mental health in adults.[71] Theories to explain the mental health benefits of water consumption are uncertain, but may be due body fluid homeostasis affecting hypothalamic regulation of stress.[72] Psychological factors associated with disease and nutrition may also contribute to mental health; for example, people with cancer may experience increased anxiety as their oral intake reduces and their physical health worsens.[73, 74] A less hydrated state may contribute to delirium through several processes, such as cerebral and renal hypoperfusion, increased concentration of medications and renal impairment.[75, 76]

It is difficult to compare our findings with previous studies reporting the associations between anxiety and hydration status, due to the use of different definitions of anxiety, and the tendency of investigators to ‘cluster’ this symptom with other factors (e.g. as restlessness and agitation.[47]). Some authors have reported associations between hydration and agitation/anxiety with mixed results; for example, Morita et al[43] found no association between clinician-rated scores of agitation for dying people with abdominal cancer, who were receiving artificial hydration compared to placebo. Similarly, Bruera et al[47] comparing outcomes for hydration vs placebo, found anxiety scores improved in the placebo group following 4 days of artificial hydration, but no difference existed at 7 days.[47] However, a study by Lokker et al[77] reported a higher frequency of agitation in dying people (cancer and non-cancer) who received higher amounts of fluid daily.

In our current study, less hydration was associated with the increased severity of dehydration (as measured by the Morita Dehydration Score), which is consistent with studies using this tool to assess physical signs of hydration.[5, 43] Oedematous participants had higher H^2^/R compared to non-oedematous participants, which suggests higher total body water. This finding is consistent with previous research;[5] however, it is not possible to determine intracellular or extracellular volumes without regression equations, which are methodologically limited in advanced cancer.[25, 26, 78, 79, 80]

Breathlessness was associated with lower H^2^/R (less hydration). However, possible explanations may relate to underlying mechanisms of breathlessness, such as the influence of peripheral and central chemoreceptors.[81, 82] However, associations between artificial hydration and breathlessness has been reported in previous studies of dying people with cancer; for example, Fritzson et al [65] reported a higher level of breathlessness for individuals receiving artificial hydration. Higher H^2^/R (suggesting more hydration) was associated with increased pain; although pain is a common in cancer, [41, 47, 83] the nature of its relationship with hydration status is unknown. Potential explanations could be due to factors related to direct effects of cancer (location and severity of the cancer affecting fluid volume homeostasis) and indirect effects, such as renal impairment and accumulation of biochemical abnormalities.

Survival was not statistically associated with H^2^/R which suggests no significant association between hydration status and prognosis. This finding differs from our previous study,[5] which demonstrated that survival was shorter in people with lower H^2^/R (i.e. less hydration). Furthermore, survival for the current cohort was shorter than the 2016 sample (median 35 days vs 62 days). Methodological differences between the studies can potentially explain differences in survival. For example, to be eligible for the 2016 study, participants were required to receive serum biochemistry tests within seven days of the first bioimpedance assessment. Therefore, participants without blood tests (for example, due to perceived lack of clinical usefulness for people with a short prognosis), were not eligible for recruitment. Participants were not required to have blood tests to participate in the current study, thus increasing the potential to recruit those who were in the ≤7 days of life.

Hydration status of dying participants did not significantly change compared to baseline assessment. This finding, together with the finding that 10.4% of all participants were ‘less hydrated’, may suggest that dehydration in the dying is not as highly prevalent as previous estimates.[84] No studies are available to directly compare our finding of a lack of association between hydration status and clinical outcomes (agitation, pain and respiratory tract secretions) in people dying with cancer. However, consistent with our finding, there is evidence that artificial hydration is not significant in causing respiratory tract secretions in the dying.[77]

### Limitations

This study describes a predominantly white UK population, in specialist palliative care units, in the last month of life. The numbers included in this analysis are small and we are not able to determine causation (of the studied variables) because the study is observational. The global COVID19 pandemic affected participant and consultee recruitment, due to restrictions on face-to-face contact, which contributed to our inability to achieve the target sample size. Furthermore, we did not achieve the target for the sub-analysis of participants in the dying phase (n=18 recruited with a target of n=30), which affects the validity of these findings. Some of the recruitment challenges were due to practical differences between research sites. For example, recruitment did not occur every day across sites, meaning some people died or were discharged prior to research assessment. Similarly, although having a process to ensure ongoing study participation following discharge, we were unable to conduct the ‘dying phase assessments’ on some participants who were discharged and then later readmitted when they were dying.

### Implications to current policy, practice and research

These findings may potentially support healthcare professionals’ decision-making regarding assessment of hydration status in people with advanced cancer, by providing evidence of the clinical outcomes related to hydration status. We do not provide guidance on the appropriate use (or non-use) of artificial hydration in advanced cancer; however, our data provides evidence about hydration assessment, to support future intervention studies evaluating the use of artificial hydration in clinical practice.

Our data demonstrates the feasibility of usings advance consent methods for palliative care research. Investigators can use the advance consent process in studies where participants are expected to lose capacity (e.g., delirium, agitation and the dying). Consequently, this method can potentially improve the evidence base (and hence, quality of care) for the dying. Future BIA/BIVA research can involve hydration assessment of specific cancers and associated variables (e.g., stage of illness, ethnicity and gender).

Furthermore, in the absence of an established core-outcome set to assess hydration in advanced disease, or data suggests that BIA/BIVA would be a potential tool to identify clinical factors related to hydration. BIA/BIVA can potentially support further research about personalised hydration.[85] Researchers can use several bioimpedance methods to conduct detailed analysis of fluid volume compartments (e.g. multifrequency bioimpedance analysis, bioimpedance spectroscopy and multi-segment bioimpedance), to examine how fluid distribution (e.g., third spacing, oedema, lymphoedema) affects clinical outcomes in cancer.[86, 87, 88, 89]

## Conclusion

In advanced cancer, hydration status was associated with specific physical signs and symptoms. No significant associations between survival and hydration status were recorded. In the dying phase, hydration status did not significantly change compared to baseline, and was not associated with symptoms. Further work can use BIA/BIVA to standardise the process to identify clinically relevant outcomes for hydration studies, to establish a core outcome set to evaluate how hydration affects symptoms and quality of life in cancer.

### Ethics

Written consent was obtained from all study participants; this included consent to report individual patient data in publication. Participant consent was recorded in a research recruitment log. All participants were provided with a copy of their consent forms. Ethics committee, North West - Haydock Ethics Research Ethics Committee, gave ethical approval for this work (REC: 17/NW/0050; IRAS ID: 196540). The study is supported by the Cancer Research UK Liverpool Cancer Trials Unit (LCTU). The LCTU is part of the Cancer Trials Research Centre which has UKCRC Clinical Trials Unit full registration, ensuring high standards of regulatory and quality control. The study was sponsored by the University of Liverpool. The researchers were independent from funders and sponsors. The sponsors and funder did not have access to the data or a role in the data analysis, results interpretation or writing the manuscript.

## Supporting information

Peripheral oedema assessment

Quality of life assessment

Performance status

The Edmonton Symptom Assessment System

Morita Dehydration Assessment Score

Normality test

Univariate analysis

Proxy measurement of comfort recorded by a healthcare professional or caregiver

## Data Availability

All data produced in the present work are contained in the manuscript

## Acknowledgements

This research was funded by, the Academy of Medical Sciences (£24,250), UKH Foundation (£5,000), Anne Duchess of Westminster Charity (£5,000) and the Liverpool Clinical Commissioning Group (CCG) Research Capability Funding (RCF) streams (£22029), Non recurrent contingency funding, National Institute for Health Research (NIHR) North West Coast Clinical Research Network, (£19,086.57). The posts of ACN and SS were funded by Marie Curie.[90] We thank Fran Westwell, Rachel Perry and Joanne Bell for assisting with the data collection.

## Supporting Information

- Dehydration Symptom Questionnaire (Appendix)
- The Edmonton Symptom Assessment System (appendix)
- Morita Dehydration Assessment Score (appendix)
- Peripheral oedema assessment (appendix)
- Quality of Life assessment (appendix)
- Performance status (appendix)
- Myoclonus assessment (appendix)
- Normality test
- Univariate analysis (appendix)
- Proxy measurement of comfort recorded by a healthcare professional or caregiver
- The Hotelling’s test of paired BIVA vector data to compare baseline and end of life care assessments

