## Supplementary material for "Non-invasive technology to assess hydration status in advanced cancer to explore relationships between fluid status and symptoms: an observational study using bioelectrical impedance analysis": Peripheral oedema assessment

Version 1: 10/10/2016

The Marie Curie
Palliative Care Institute
LIVERPOOL

**Oedema assessment tool**

| | |
|---|---|
| **UPPER LIMBS** | |
| **LEFT ARM** | **RIGHT ARM** |
| ☐ [0] No oedema<br>☐ [1] oedema | ☐ [0] No oedema<br>☐ [1] oedema |
| **LOWER LIMBS** | |
| **LEFT LEG** | **RIGHT LEG** |
| ☐ [0] No oedema<br>☐ [1] oedema | ☐ [0] No oedema<br>☐ [1] oedema |
| **TORSO** | |
| **THORAX (from radiology)** | **ABDOMEN** |
| ☐ [0] No pleural effusion<br>☐ [1] Pleural effusion present | ☐ [0] No ascites<br>☐ [1] ascites present |

Total score = (maximum 6)
