## Supplementary material for "Non-invasive technology to assess hydration status in advanced cancer to explore relationships between fluid status and symptoms: an observational study using bioelectrical impedance analysis": Quality of life assessment

**Version 1: 10/10/2016**

The Marie Curie
Palliative Care Institute
LIVERPOOL

**FACT-G7 (Version 4)**

Below is a list of statements that other people with your illness have said are important**. Please circle or mark one number per line to indicate your response as it applies to the past 7 days.**

| | | Not at all | A little bit | Some-what | Quite a bit | Very much |
|---|---|---|---|---|---|---|
| GP1 | I have a lack of energy | 0 | 1 | 2 | 3 | 4 |
| GP4 | I have pain | 0 | 1 | 2 | 3 | 4 |
| GP2 | I have nausea | 0 | 1 | 2 | 3 | 4 |
| GE6 | I worry that my condition will get worse | 0 | 1 | 2 | 3 | 4 |
| GF5 | I am sleeping well | 0 | 1 | 2 | 3 | 4 |
| GF3 | I am able to enjoy life | 0 | 1 | 2 | 3 | 4 |
| GF7 | I am content with the quality of my life right now | 0 | 1 | 2 | 3 | 4 |
