## Supplementary material for "Non-invasive technology to assess hydration status in advanced cancer to explore relationships between fluid status and symptoms: an observational study using bioelectrical impedance analysis": Performance status

The Marie Curie
Palliative Care Institute
LIVERPOOL

**Eastern Cooperative Oncology Group (ECOG) classification of performance status[1]**

| ECOG | Description |
|---|---|
| 0 | Fully active, able to carry on all pre-disease performance without restriction. |
| 1 | Restricted in physically strenuous activity but ambulatory and able to carry out work of a light or sedentary nature, e.g., light house work, office work. |
| 2 | Ambulatory and capable of all selfcare but unable to carry out any work activities. Up and about more than 50% of waking hours. |
| 3 | Capable of only limited selfcare, confined to bed or chair more than 50% of waking hours. |
| 4 | Completely disabled. Cannot carry on selfcare. Totally confined to bed or chair |

1. Oken MM, Creech RH, Tormey DC, et al. Toxicity and response criteria of the Eastern Cooperative Oncology Group. Am J Clin Oncol 1982;**5**(6):649-55.
