## Supplementary material for "Non-invasive technology to assess hydration status in advanced cancer to explore relationships between fluid status and symptoms: an observational study using bioelectrical impedance analysis": The Edmonton Symptom Assessment System

Alberta Health Services

Covenant Health

### Edmonton Symptom Assessment System:
**(revised version) (ESAS-R)**

**Please circle the number that best describes how you feel NOW:**

| | | | | | | | | | | | | |
|---|---|---|---|---|---|---|---|---|---|---|---|---|
| No Pain | **0** | **1** | **2** | **3** | **4** | **5** | **6** | **7** | **8** | **9** | **10** | Worst Possible Pain |
| No Tiredness *(Tiredness = lack of energy)* | **0** | **1** | **2** | **3** | **4** | **5** | **6** | **7** | **8** | **9** | **10** | Worst Possible Tiredness |
| No Drowsiness *(Drowsiness = feeling sleepy)* | **0** | **1** | **2** | **3** | **4** | **5** | **6** | **7** | **8** | **9** | **10** | Worst Possible Drowsiness |
| No Nausea | **0** | **1** | **2** | **3** | **4** | **5** | **6** | **7** | **8** | **9** | **10** | Worst Possible Nausea |
| No Lack of Appetite | **0** | **1** | **2** | **3** | **4** | **5** | **6** | **7** | **8** | **9** | **10** | Worst Possible Lack of Appetite |
| No Shortness of Breath | **0** | **1** | **2** | **3** | **4** | **5** | **6** | **7** | **8** | **9** | **10** | Worst Possible Shortness of Breath |
| No Depression *(Depression = feeling sad)* | **0** | **1** | **2** | **3** | **4** | **5** | **6** | **7** | **8** | **9** | **10** | Worst Possible Depression |
| No Anxiety *(Anxiety = feeling nervous)* | **0** | **1** | **2** | **3** | **4** | **5** | **6** | **7** | **8** | **9** | **10** | Worst Possible Anxiety |
| Best Wellbeing *(Wellbeing = how you feel overall)* | **0** | **1** | **2** | **3** | **4** | **5** | **6** | **7** | **8** | **9** | **10** | Worst Possible Wellbeing |
| No ___________ Other Problem *(for example constipation)* | **0** | **1** | **2** | **3** | **4** | **5** | **6** | **7** | **8** | **9** | **10** | Worst Possible ___________ |

Patient's Name ______________________________

Date ________________ Time ________________

Completed by (check one):

- ☐ Patient
- ☐ Family caregiver
- ☐ Health care professional caregiver
- ☐ Caregiver-assisted

**BODY DIAGRAM ON REVERSE SIDE**

**Please mark on these pictures where it is that you hurt:**

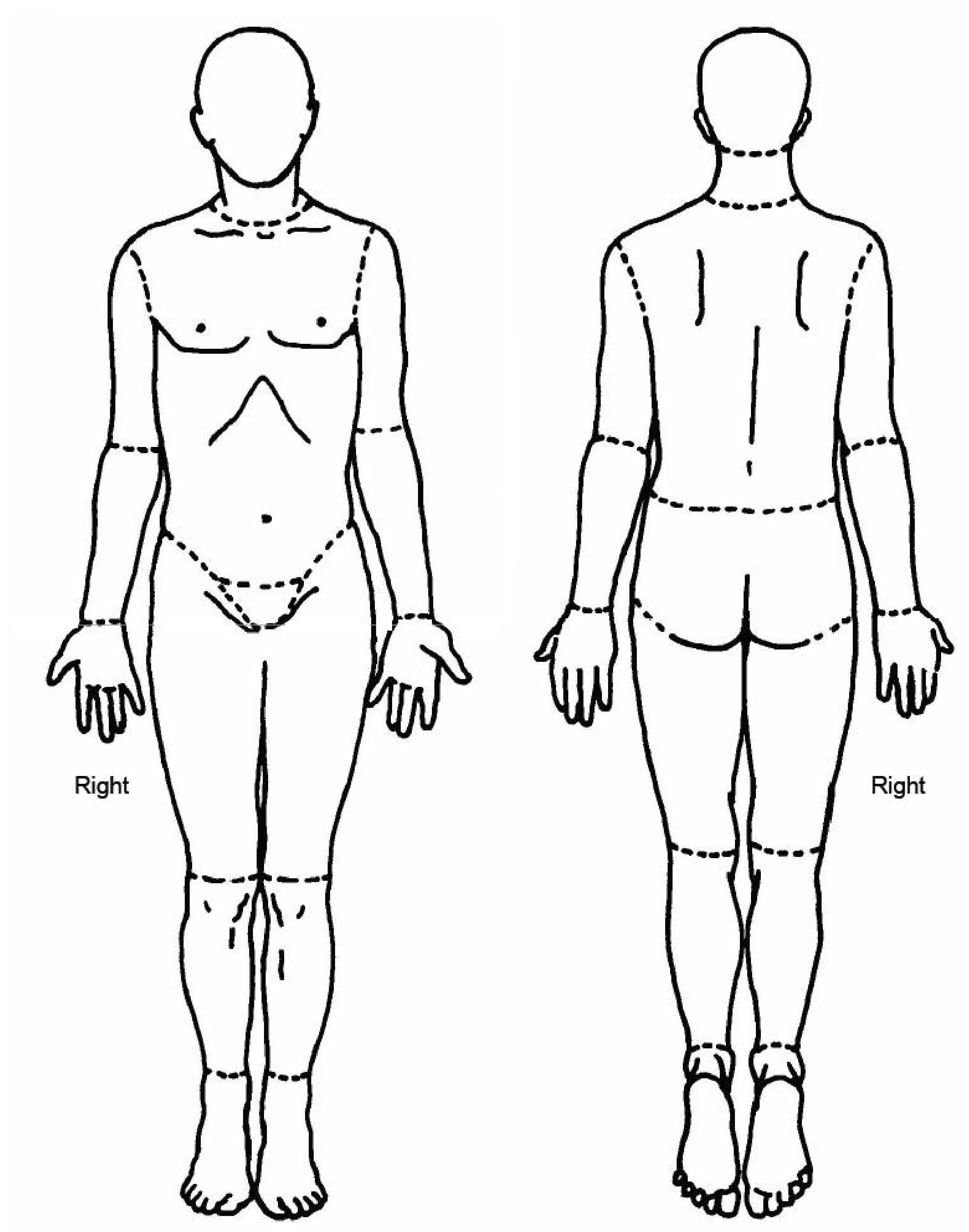
