## Supplementary material for "Non-invasive technology to assess hydration status in advanced cancer to explore relationships between fluid status and symptoms: an observational study using bioelectrical impedance analysis": Morita Dehydration Assessment Score

The Marie Curie
Palliative Care Institute
LIVERPOOL

### Hydration questionnaire (adapted from Morita 2006)

**Assessment type**

**Baseline** ☐ **Follow up** ☐

**DATE:**

**Clinical Examination guidance**

For each patient indicate a score for each variable and then calculate the total hydration score:

| | | **SCORE** | |
|---|---|---|---|
| **VARIABLE** | **0** | **1** | **2** |
| Moisture of mucous membranes of mouth | Moist ☐ Moist oral mucous membrane **AND** moist tongue palpation. | Somewhat dry ☐ **EITHER** dry oral mucous membrane with moist tongue **OR** moist mucous membrane with dry tongue on palpation. | Dry ☐ Dry oral mucous membrane **PLUS** dry tongue on palpation, with or without presence of tongue furrows, coated tongue or oral/mucous lesions |
| Moisture of the axilla | Moist ☐ Moisture present on palpation | Dry ☐ Dry on palpation | |
| Sunkenness of eyes | Normal ☐ Both eyes appear normal without recess in the socket | Slightly sunken ☐ Both eyes appear slightly recessed in socket. | Sunken ☐ Both eyes appear deeply/completely recessed in socket. |

**Total = /5**
