## Supplementary material for "Non-invasive technology to assess hydration status in advanced cancer to explore relationships between fluid status and symptoms: an observational study using bioelectrical impedance analysis": Normality test

**Normality test for the regression analysis**

The Shapiro-Wilk test showed that the distribution of the impedance index departed significantly from normality (W = 0.89, p <0.01).

**Tests of Normality**

| | Kolmogorov-Smirnov[a] | | | Shapiro-Wilk | | |
|---|---|---|---|---|---|---|
| | Statistic | df | Sig. | Statistic | df | Sig. |
| Impedance index | .119 | 124 | <.001 | .885 | 124 | <.001 |

*. This is a lower bound of the true significance.

a. Lilliefors Significance Correction

Based on this outcome, we converted the impedance index to $\log^{10}$ values and repeated the Shapiro-Wilk test, which did not show evidence of non normality (W = 0.98, p= 0.053).

**Tests of Normality**

| | Kolmogorov-Smirnov[a] | | | Shapiro-Wilk | | |
|---|---|---|---|---|---|---|
| | Statistic | df | Sig. | Statistic | df | Sig. |
| Lg10_impedance_index | .056 | 124 | .200* | .979 | 124 | .053 |

*. This is a lower bound of the true significance.

a. Lilliefors Significance Correction

We visually examined the histogram of the impedance index and the QQ plot and we were satisfied that the $\log^{10}$ impedance index data met the normality requirements for the backward regression analysis.

Non-invasive technology to assess hydration status in advanced cancer to explore relationships between fluid-status and symptoms at the end-of-life: an observational study using bioelectrical impedance analysis

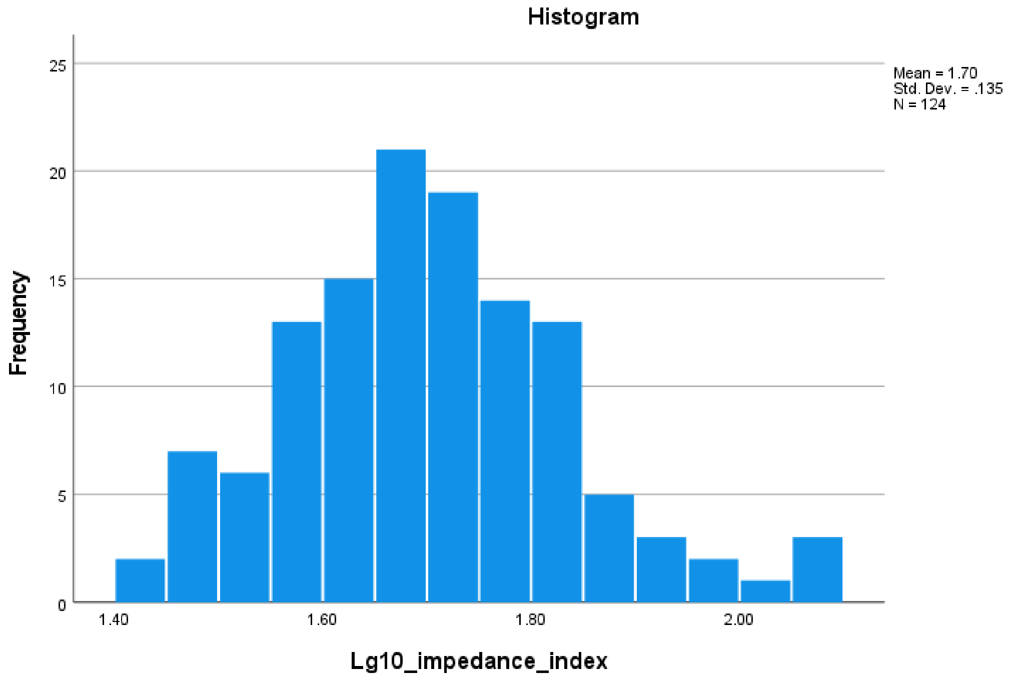


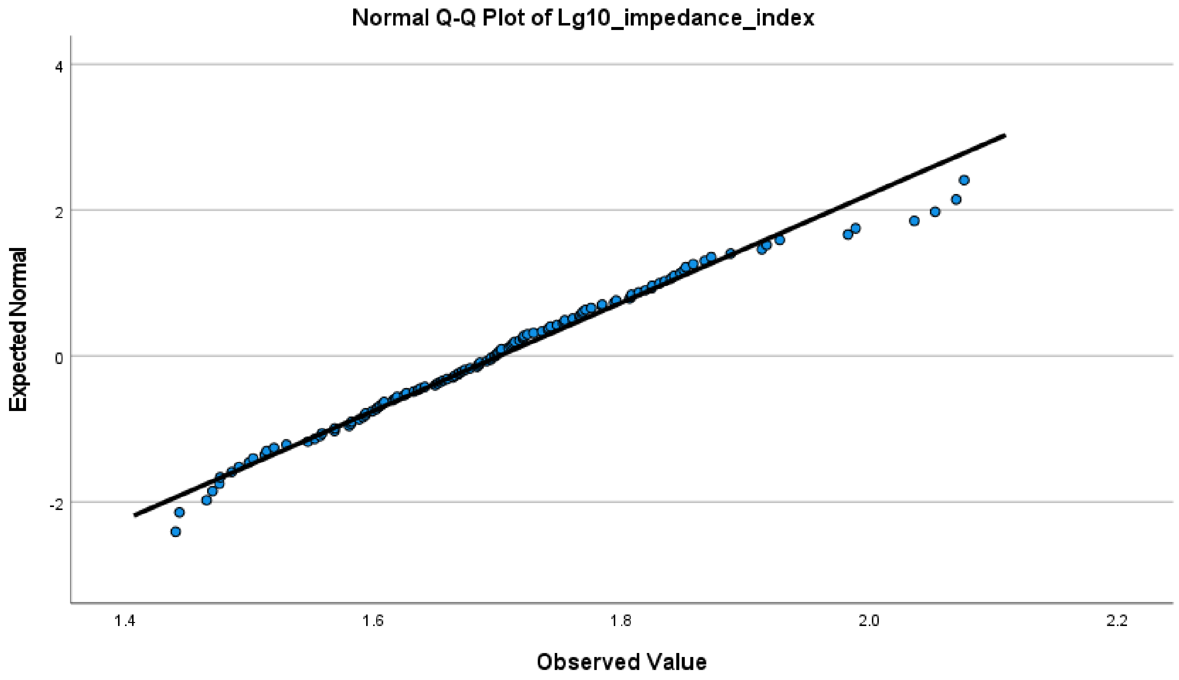

Non-invasive technology to assess hydration status in advanced cancer to explore relationships between fluid-status and symptoms at the end-of-life: an observational study using bioelectrical impedance analysis

**Test of normality of repeat impedance assessment (i.e. end of life, dying phase assessment)**

**Tests of Normality**

| | Kolmogorov-Smirnov[a] | | | Shapiro-Wilk | | |
|---|---|---|---|---|---|---|
| | Statistic | df | Sig. | Statistic | df | Sig. |
| Follow up impedance index | .204 | 18 | .046 | .915 | 18 | .105 |

a. Lilliefors Significance Correction

The Shapiro-Wilk test showed that the distribution of the impedance index did not significantly depart from normality (W = 0.92, p =0.105). Therefore, we used the unadjusted follow-up impedance index in the assessment of repeated values as outlined in the methods.
