## Supplementary material for "Non-invasive technology to assess hydration status in advanced cancer to explore relationships between fluid status and symptoms: an observational study using bioelectrical impedance analysis": Univariate analysis

### Univariate analysis of association between impedance index and other factors (Spearman rank)

| | | |
|---|---|---|
| Impedance index | Correlation Coefficient | 1.000 |
| | Sig. (2-tailed) | . |
| | N | 124 |
| Sex of patient | Correlation Coefficient | -.378** |
| | Sig. (2-tailed) | <.001 |
| | N | 124 |
| Age of patient at recruitment | Correlation Coefficient | -.101 |
| | Sig. (2-tailed) | .262 |
| | N | 124 |
| WHO performance status | Correlation Coefficient | -.151 |
| | Sig. (2-tailed) | .094 |
| | N | 124 |
| Edmonton, current pain assessment | Correlation Coefficient | -.009 |
| | Sig. (2-tailed) | .924 |
| | N | 124 |
| Edmonton, current tiredness assessment | Correlation Coefficient | -.120 |
| | Sig. (2-tailed) | .185 |
| | N | 124 |
| Edmonton, current drowsiness assessment | Correlation Coefficient | -.096 |
| | Sig. (2-tailed) | .291 |
| | N | 124 |
| Edmonton, current nausea assessment | Correlation Coefficient | -.065 |
| | Sig. (2-tailed) | .472 |
| | N | 124 |
| Edmonton, current appetite assessment | Correlation Coefficient | -.273** |
| | Sig. (2-tailed) | .002 |
| | N | 124 |
| Edmonton, current shortness of breath assessment | Correlation Coefficient | -.111 |
| | Sig. (2-tailed) | .221 |
| | N | 124 |
| Edmonton, current depression assessment | Correlation Coefficient | -.159 |
| | Sig. (2-tailed) | .078 |
| | N | 124 |
| | Correlation Coefficient | -.192* |

| | | |
|---|---|---|
| Edmonton, current anxiety assessment | Sig. (2-tailed) | .032 |
| | N | 124 |
| Edmonton, current wellbeing assessment | Correlation Coefficient | -.126 |
| | Sig. (2-tailed) | .162 |
| | N | 124 |
| Burge, current thirst | Correlation Coefficient | -.005 |
| | Sig. (2-tailed) | .959 |
| | N | 124 |
| Burge, current dry mouth | Correlation Coefficient | -.171 |
| | Sig. (2-tailed) | .057 |
| | N | 124 |
| Burge, current taste in mouth | Correlation Coefficient | -.120 |
| | Sig. (2-tailed) | .184 |
| | N | 124 |
| Morita, moisture of mucous membranes in mouth | Correlation Coefficient | -.242** |
| | Sig. (2-tailed) | .007 |
| | N | 124 |
| Morita, moisture of the axilla | Correlation Coefficient | -.042 |
| | Sig. (2-tailed) | .644 |
| | N | 124 |
| Morita, sunkenness of eyes | Correlation Coefficient | -.333** |
| | Sig. (2-tailed) | <.001 |
| | N | 124 |
| Myoclonus present? | Correlation Coefficient | .139 |
| | Sig. (2-tailed) | .124 |
| | N | 124 |
| Oedema present? | Correlation Coefficient | .509** |
| | Sig. (2-tailed) | <.001 |
| | N | 124 |
| 'I have a lack of energy'. Scored over 7 days | Correlation Coefficient | -.141 |
| | Sig. (2-tailed) | .118 |
| | N | 124 |
| 'I have pain'. Scored over 7 days | Correlation Coefficient | -.118 |
| | Sig. (2-tailed) | .191 |
| | N | 124 |
| 'I have nausea'. Scored over 7 days | Correlation Coefficient | -.085 |
| | Sig. (2-tailed) | .347 |
| | N | 124 |
| 'I worry that my condition will get worse'. Scored over 7 days | Correlation Coefficient | -.190* |
| | Sig. (2-tailed) | .034 |

| | | |
|---|---|---|
| | N | 124 |
| 'I am sleeping well'. Scored over 7 days | Correlation Coefficient | -.235** |
| | Sig. (2-tailed) | .009 |
| | N | 124 |
| 'I am able to enjoy life'. Scored over 7 days | Correlation Coefficient | .083 |
| | Sig. (2-tailed) | .361 |
| | N | 124 |
| 'I am content with the quality of my life right now'. Scored over 7 days | Correlation Coefficient | .150 |
| | Sig. (2-tailed) | .096 |
| | N | 124 |
