## Supplementary material for "Non-invasive technology to assess hydration status in advanced cancer to explore relationships between fluid status and symptoms: an observational study using bioelectrical impedance analysis": Proxy measurement of comfort recorded by a healthcare professional or caregiver

The Marie Curie
Palliative Care Institute
LIVERPOOL

**Proxy comfort score**

### DATE:

Please circle the number that best describes how comfortable the participant is.

**1. AGITATION**

| | | | | | | |
|---|---|---|---|---|---|---|
| Not at all agitated | **0** | **1** | **2** | **3** | **4** | Extremely agitated |

**2. PAIN**

| | | | | | | |
|---|---|---|---|---|---|---|
| Not in pain | **0** | **1** | **2** | **3** | **4** | severe pain |

**3. RESPIRIATORY TRACT SECRETIONS**

| | | | | | | |
|---|---|---|---|---|---|---|
| No respiratory tract secretions audible | **0** | **1** | **2** | **3** | **4** | Extremely loud secretions |
