## Supplemental Data 1 for "Non-invasive technology to assess hydration status in advanced cancer to explore relationships between fluid status and symptoms: an observational study using bioelectrical impedance analysis"

### Hotelling’s test of paired data

#### Hotelling’s test of paired bioimpedance data (baseline and dying phase assessments) for all patients (n = 18)

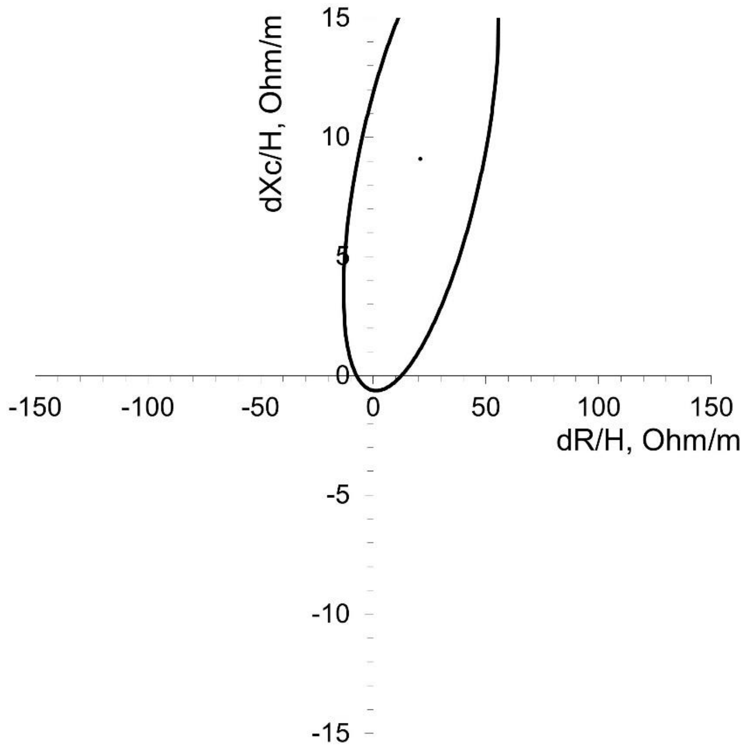


#### Hotelling’s test of paired bioimpedance data (baseline and dying phase assessments) for females (n = 9)

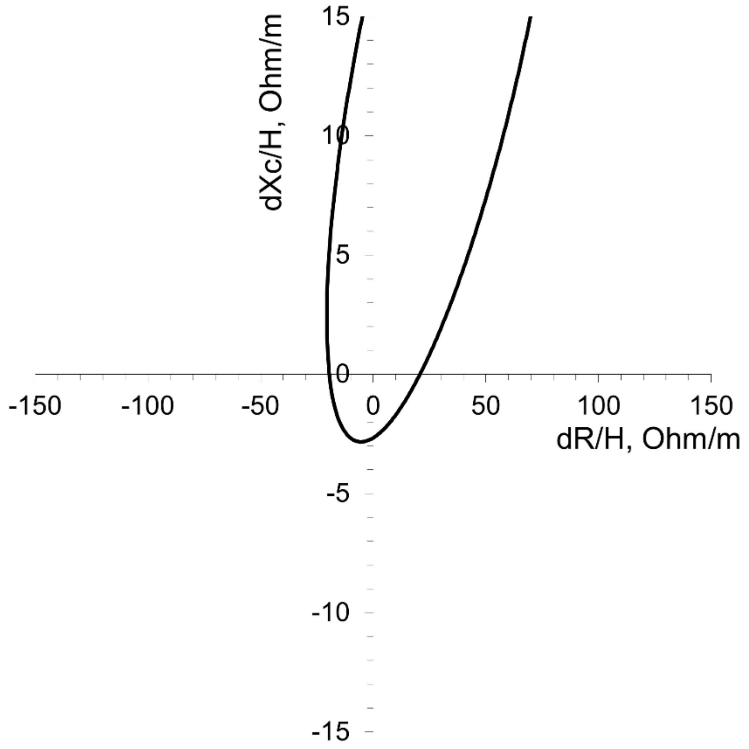


**Hotelling's test of paired bioimpedance data (baseline and dying phase assessments) for males (n = 9)**

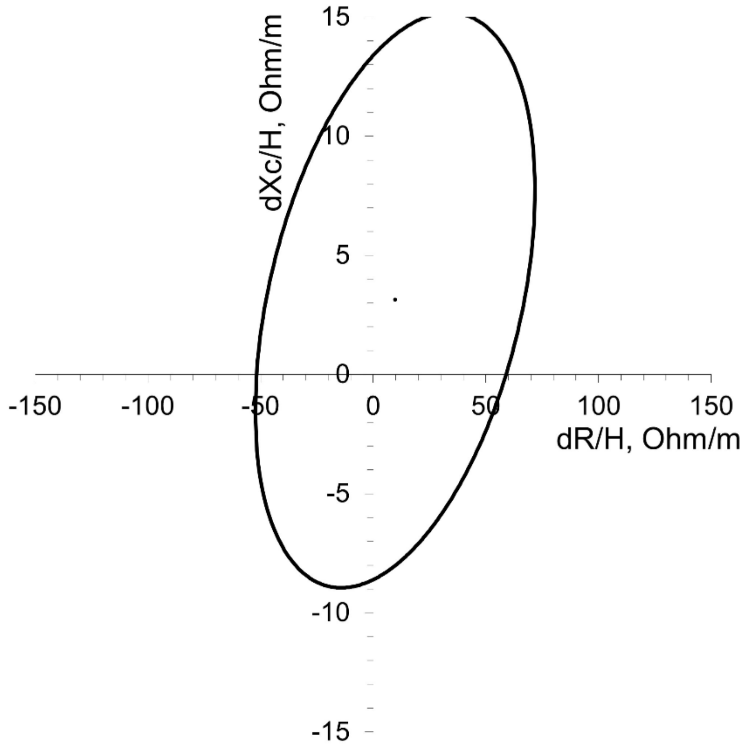
